# Beyond Black vs White: Racial/Ethnic Disparities in Chronic Pain including Hispanic, Asian, Native American, and Multiracial U.S. Adults

**DOI:** 10.1101/2021.08.10.21261852

**Authors:** Anna Zajacova, Hanna Grol-Prokopczyk, Roger Fillingim

**Affiliations:** University of Western Ontario; University at Buffalo, State University of New York; University of Florida

## Abstract

Previous literature on race/ethnicity and pain has rarely included all major U.S. racial groups or examined the sensitivity of findings to different pain operationalizations. Using data from the 2010-2018 National Health Interview Surveys on adults 18+ (N=273,972), we calculate the weighted prevalence of six definitions of pain to provide a detailed description of chronic pain in White, Black, Hispanic, Asian, Native American, and multiracial groups. We also estimate logistic models to obtain relative disparities, including net of demographic and socioeconomic (SES) factors; finally, we calculate average predicted probabilities to show prevalence disparities in absolute terms. We find that Asian Americans have the lowest pain prevalence across all pain definitions or model specifications. In contrast, Native American and multiracial adults have the highest pain prevalence. This pain excess is due to the lower SES among Native Americans but remains significant and unexplained among multiracial adults. Whites, Blacks, and Hispanics fall in between. In this trio, Hispanics have the lowest prevalence, an advantage not attributable to immigrant status or SES. While most prior research focused on Black-White comparisons, these two groups differ relatively little. Blacks report lower prevalence of less severe pain definitions than Whites, but higher prevalence of severe pain. Net of SES, however, Blacks have significantly lower pain across all definitions. Overall, racial disparities are larger than previously recognized once all major racial groups are included, and these disparities are largely consistent across different operationalizations of pain.

## IN BRIEF

U.S. racial pain disparities are larger than previously recognized; pain prevalence is strikingly high among Native American and multiracial adults and lowest among Asian Americans.

Despite the existence of a large literature on race/ethnicity and pain, important fundamental facts regarding chronic pain disparities in the general adult U.S. population have not been established. In part, this reflects the focus of much prior research on Black-versus-White comparisons [6; 13]. However, even this narrowly-defined comparison has yielded somewhat inconsistent results: some studies find no difference between Blacks and Whites [19]; some find higher prevalence of pain among Blacks [23; 25; 38; 50]; others find the opposite [22; 55; 57; 58]; and some studies report both patterns within one analysis, depending on the specific pain measure used [11; 16; 33; 34; 42]. Further, although Hispanics now constitute the largest non-White group, comprising over 21% of the total U.S. population [48], limited attention has been given to their experiences of pain. Even rarer are studies focused on Asian American and Native American adults. Finally, to the best of our knowledge, pain in multiracial Americans has never been systematically studied. This is troubling given that the multiracial population is the fastest-growing racial subgroup in the U.S. and is projected to more than double between 2020 to 2050, from 9.5 to 20.5 million [47]. Additionally, prior work on racial/ethnic pain disparities has rarely tested whether or how estimates vary across different pain operationalizations. This is a critical gap given not only the inconsistent results for the Black-White disparities cited above, but also considering that the pain definitions across epidemiological studies of chronic pain are “highly inconsistent, with virtually no two studies … using the exact same criteria” [45, p. 2092]. The existing research gaps thus neglect major segments of the U.S. population and potential differences in disparity patterns across pain severity levels.

This study makes three primary contributions to our understanding of U.S. racial/ethnic differences in chronic pain. First, we use large, up-to-date, nationally representative data to characterize pain prevalence not only among Whites and Blacks, as the most studied and compared groups, but also among Hispanics, Asian Americans, Native Americans, and multiracial adults, as well as specific ‘main racial background’ mentioned by multiracial adults. Second, we analyze whether the observed racial/ethnic differences are explained by key demographic and socioeconomic factors known to impact chronic pain at the population level, such as immigrant status, education, and family income [19; 20; 57]. And third, we present pain prevalence estimates for six commonly used pain operationalizations: severe pain, high-impact chronic pain, widespread pain, persistent pain over 3 months, persistent pain over 6 months, and ‘any pain.’ This enables us to document racial/ethnic disparities at differing levels of pain severity, to potentially reconcile the complex comparisons between White and Black adults, and, generally, to clarify whether different pain measures lead to different conclusions regarding racial/ethnic disparities.

Overall, these three contributions generate a more complete picture of U.S. racial/ethnic disparities across a range of widely-used pain definitions. We identify groups at particularly high risk of chronic pain, highlighting potential unmet needs for pain prevention and treatment efforts. As well, we identify groups at relatively low risk, inviting further exploration of protective characteristics.

## METHODS

### Data

The analyses use National Health Interview Survey (NHIS) data from 2010-2018. The NHIS is a large, repeated cross-sectional survey representative of the U.S. non-institutionalized population. It is one of the main sources of information about the health of U.S. adults. The years were selected to maximize sample size and yield up-to-date information while minimizing changes in sampling design and questionnaire wording. The year 2010 is when the NHIS began asking global pain questions, while 2018 is the most recent year data was available at the time of the writing, as well the last year before a major redesign in 2019, which could affect comparability to prior years. We used NHIS data harmonized by Integrated Public Use Microdata Series (IPUMS, https://ipums.org/), which is the optimal version for analyzing NHIS data from multiple years [44]. The data are publicly available and de-identified; as such, this analysis is exempt from IRB approvals.

### The target population

comprises community-dwelling adults age 18 and older. Proxy responses (fewer than 0.5 percent NHIS sample adults) were excluded as pain responses are inherently subjective; thus, a proxy’s reports cannot be substituted for self-reports. Sample sizes, which range from N=57,518 for high-impact chronic pain to 273,972 for “any pain” and widespread pain, are shown in Table 1; additional details on their derivation is in the Supplement. Briefly, for all 6 pain definitions, over 99% of respondents provided valid answers to necessary pain and/or race questions; thus, fewer than 1% were excluded from analyses.

**Table 1.** The six pain definitions, sample sizes, years when available, wording of questions, and coding. NHIS 2010-2018.

| Pain definition and sample size | Years | Question wording | Coding (all dichotomized) | Original NHIS variables |
| --- | --- | --- | --- | --- |
| Severe pain<br>(N=103,349) | 2010-2015<br>and 2018 | Level of pain, last time had pain: a little, a lot, or somewhere in between (also uses painfreq3mo item) | "A lot of pain," on most or every day (using painfreq3mo)= 1; otherwise 0 | painfeelevl |
| High-impact pain<br>(N=57,518) | 2016, 2017 | How often did pain limit life or work activities: never, some days, most days, every day | Never or some days=0<br>Most or every day =1 | painlmt6mo |
| Widespread pain<br>(N=273,972) | 2010-2018 | No specific questions, based on items for 'any pain' | Positive response to 3 or more sites with pain=1; otherwise 0 | See 'any pain' below |
| Persistent (3 mos)<br>(N=103,439) | 2010-2015<br>and 2018 | Frequency of pain in last 3 months: never, some days, most days, every day | Never or some days=0<br>Most or every day =1 | painfreq3mo |
| Persistent (6 mos)<br>(N=57,541) | 2016, 2017 | Frequency of pain in last 6 months: never, some days, most days, every day | Never or some days=0<br>Most or every day =1 | painfreq6mo |
| Any pain<br>(N=273,972) | 2010-2018 | During the past 3 months, did you have [low back, neck, facial/jaw, headache/migraine, joint] pain? | Positive response to either site=1;<br>negative response to all =0 | facepain3mo lbpain3mo migrain3mo<br>neckpain3mo joint3mo |

### Variables

#### We analyze six pain measures

severe pain, high-impact chronic pain, widespread pain, persistent pain over 3 months, persistent pain over 6 months, and any pain. Table 1 summarizes the wording of questions, years of collection, and coding for all measures.

“Any pain” is constructed from questions about pain in five anatomical sites, chosen for being the most common and/or disabling types [41]. These were posed in each year from 2010-2018 to all sample adults. Four questions were of the form: “During the past three months, did you have [low back pain, neck pain, severe headache or migraine, or facial or jaw ache or pain]?” The fifth site (joint pain) was collected with two items. Respondents were asked whether they had “any symptoms of pain, aching, or stiffness in or around a joint” and if they did, whether the onset was at least 3 months prior. We used a positive response to this follow-up question as an indicator of chronic joint pain so that all pain measures have a 3-month horizon. Respondents who indicated pain in at least one site are coded as having ‘any pain,’ following precedent [55; 57], while respondents who indicated pain in at least three sites are categorized as having widespread pain.

Since 2010, a subset or all respondents were also asked, “In the past 3 months, how often did you have pain? Would you say never, some days, most days, or every day?” Following established precedent, we dichotomized this measure as never or some days versus most or every day [33; 53]. This measure is referred to as chronic or persistent pain (3 months). Those who said they had pain on at least some days were also asked “Thinking about the last time you had pain, how much pain did you have? Would you say a little, a lot, or somewhere in between?” We used these two questions to construct a measure of severe pain, which is “a lot” of pain on most or every day.

In 2016 and 2017, the NHIS included a pain frequency item with respect to the last 6 months, worded similarly to the 3-month persistent pain item. Adults who had pain on at least some days were then asked how often the pain limited their life or work activities. Leaning on precedent, we dichotomized the answers as never or some days versus most or every day, where the latter operationalized high-impact chronic pain [11; 58].

#### The key independent variable is race/ethnicity

This variable, which combines racial and Hispanic-ethnicity self-identifications, was generated from two questions. First, respondents self-identified as Hispanic or not Hispanic (variable hispyn). The second racial self-identification (variable racenew) follows the post-1997 Office of Management and Budget standard; respondents are categorized as White, Black/African American, American Indian/Alaskan Native (AIAN), Asian, or Multiple Race. We combined these two variables into a single racial/ethnic categorization, following prior empirical precedent [26] and theoretical justification [52]. The variable is thus coded as White (reference in regression models), Black, Hispanic, Asian, Native American (or AIAN: American Indian/Alaskan Native), and multiracial. All groups other than Hispanic are non-Hispanic; we omit the adjective for parsimony.

#### Covariates include demographic and socioeconomic characteristics

Year of interview is included in all models as a continuous covariate centered on 2017. Age is in single years. Because of nonlinearities in the age-pain association [7; 16], age is centered on 60 and a quadratic term is added to capture the plateauing or declining of pain at older ages. Gender is included with male as reference. Nativity status is captured with a 3-level variable, which categorizes respondents as U.S.-born, foreign-born but in U.S. for 15 or more years, and foreign-born and in the U.S. for less than 15 years. Language of interview is dichotomized as English or other language. Marital status is coded as married, previously married, and never married; and the number of children that reside with a respondent is coded as no children, one child, and two or more children. Finally, we include three measures of socioeconomic status. Education is categorized as less than a high school diploma, high school diploma, some college, associate degree, bachelor‘s degree, and master’s or more advanced degree. GED recipients are included with the lowest education category based on prior work [54]. Family income is categorized as $0-34,999, $35,000-74,999, $75,000-99,999, and $100,000 or more. The last measure is home ownership (owns home, rents, or other arrangement), as a proxy for wealth and long-term financial wellbeing and stability [46].

### Missingness

The quality of data collected by the NHIS is high, with low missingness. As described in the Supplement, 99.2% to 99.9% of participants who were asked the race and various pain questions provided valid answers. Missingness on covariates was generally also low. Age, gender, year of interview, region of residence, and children had no missing values. Language of interview was missing for 0.004% (12 cases) out of 273,972 total cases, and home ownership was missing for 0.2%; we included the missing cases with “other” levels. Marital status, immigrant status, and education were missing in 0.2-0.4% of cases, and only income had higher proportion of missing values, at 6.8%. For these four variables, we include a “DK” category in all regression models, although findings are very similar for a complete-case analysis.

### Approach

We first estimate the weighted prevalence of each of the six pain definitions in all racial/ethnic categories (Table 2). We also test whether the prevalence differs significantly by race/ethnicity using chi-squared tests with correction for sampling weights and complex sample design [36]. We then conduct the same analytic steps separately for men and women (Supplement Table S1); as well as by the main racial background listed by multiracial adults for further understanding of this complex category (Supplement Table S2). We also list the number of respondents of each race/ethnicity in each analysis to show that all racial/ethnic categories include a sufficient number of respondents for reliable estimates (Supplement Table S3). The third descriptive step summarizes the distribution of all covariates in the total sample and for the six racial/ethnic groups, and tests for differences in their distribution by race/ethnicity using the same approach as above (Table 3).

**Table 2.**
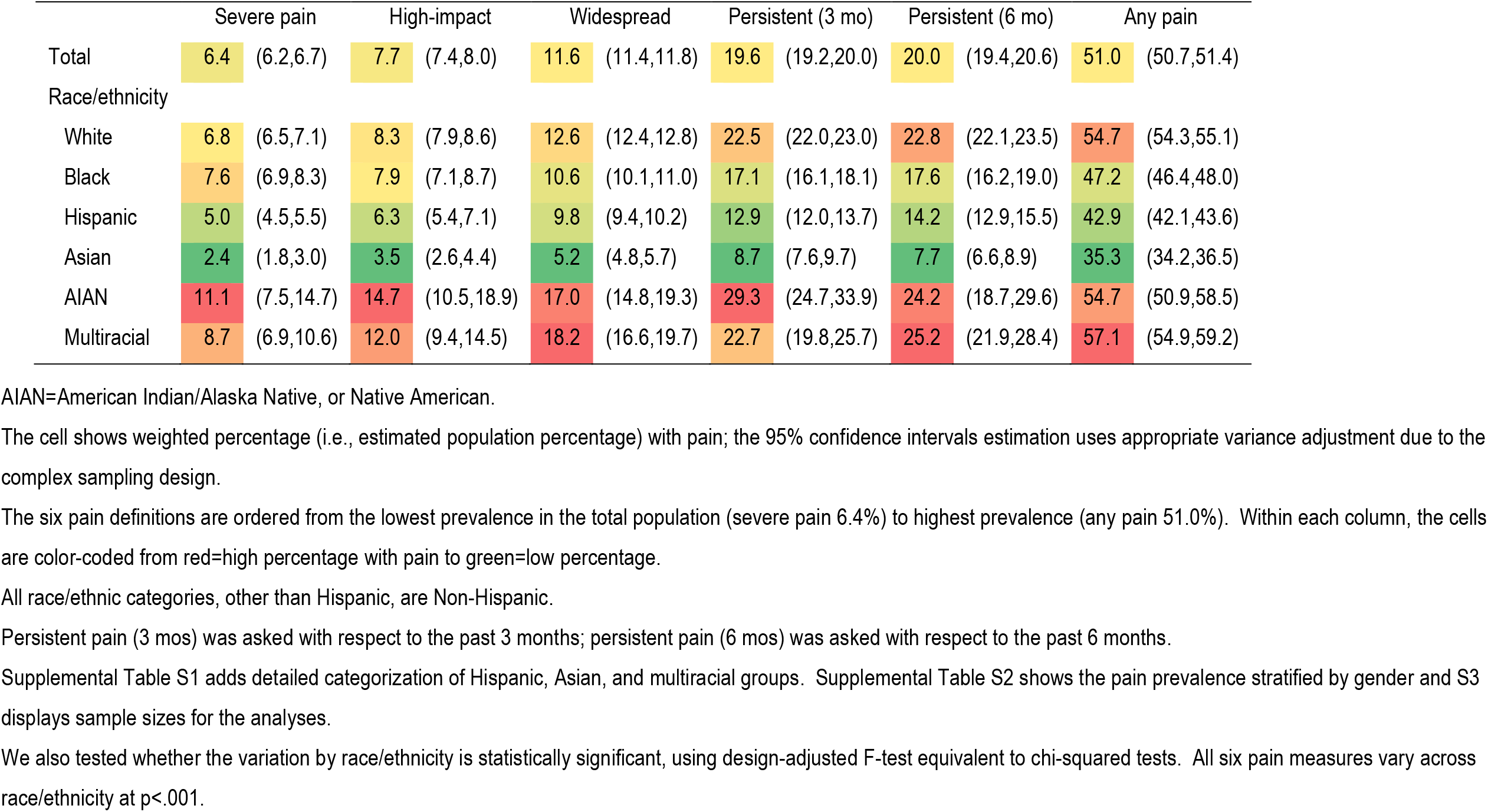
Population prevalence of pain (weighted percent and 95% CI) for six pain definitions, by race/ethnic categories, US adults age 18+.

|  | Severe pain |  | High-impact |  | Widespread |  | Persistent (3 mo) |  | Persistent (6 mo) |  | Any pain |  |
| --- | --- | --- | --- | --- | --- | --- | --- | --- | --- | --- | --- | --- |
| Total | 6.4 | (6.2,6.7) | 7.7 | (7.4,8.0) | 11.6 | (11.4,11.8) | 19.6 | (19.2,20.0) | 20.0 | (19.4,20.6) | 51.0 | (50.7,51.4) |
| Race/ethnicity |  |  |  |  |  |  |  |  |  |  |  |  |
| White | 6.8 | (6.5,7.1) | 8.3 | (7.9,8.6) | 12.6 | (12.4,12.8) | 22.5 | (22.0,23.0) | 22.8 | (22.1,23.5) | 54.7 | (54.3,55.1) |
| Black | 7.6 | (6.9,8.3) | 7.9 | (7.1,8.7) | 10.6 | (10.1,11.0) | 17.1 | (16.1,18.1) | 17.6 | (16.2,19.0) | 47.2 | (46.4,48.0) |
| Hispanic | 5.0 | (4.5,5.5) | 6.3 | (5.4,7.1) | 9.8 | (9.4,10.2) | 12.9 | (12.0,13.7) | 14.2 | (12.9,15.5) | 42.9 | (42.1,43.6) |
| Asian | 2.4 | (1.8,3.0) | 3.5 | (2.6,4.4) | 5.2 | (4.8,5.7) | 8.7 | (7.6,9.7) | 7.7 | (6.6,8.9) | 35.3 | (34.2,36.5) |
| AIAN | 11.1 | (7.5,14.7) | 14.7 | (10.5,18.9) | 17.0 | (14.8,19.3) | 29.3 | (24.7,33.9) | 24.2 | (18.7,29.6) | 54.7 | (50.9,58.5) |
| Multiracial | 8.7 | (6.9,10.6) | 12.0 | (9.4,14.5) | 18.2 | (16.6,19.7) | 22.7 | (19.8,25.7) | 25.2 | (21.9,28.4) | 57.1 | (54.9,59.2) |

**Table 3.** Characteristics of the target population in aggregate and by race/ethnic group (NHIS 2010-2018, US adults 18 and older)

|  | Total | White | Black | Hispanic | Asian | AIAN | Multi-racial |
| --- | --- | --- | --- | --- | --- | --- | --- |
| Age (mean, s.d.) | 46.8 | 49.0 | 44.3 | 40.8 | 44.4 | 44.5 | 40.4 |
| Female | 51.8 | 51.5 | 55.2 | 49.7 | 53.1 | 55.1 | 52.9 |
| Region |  |  |  |  |  |  |  |
| Northeast | 17.7 | 19.0 | 16.0 | 13.6 | 20.3 | 5.5 | 10.8 |
| North Central | 22.8 | 27.9 | 16.9 | 9.2 | 12.9 | 15.9 | 21.4 |
| South | 36.3 | 33.4 | 59.0 | 37.0 | 22.8 | 33.9 | 33.5 |
| West | 23.2 | 19.7 | 8.1 | 40.2 | 43.9 | 44.7 | 34.3 |
| Nativity |  |  |  |  |  |  |  |
| US-born | 81.9 | 94.7 | 87.0 | 42.9 | 22.1 | 93.7 | 92.9 |
| In US 15+ years | 11.7 | 3.8 | 7.5 | 37.3 | 46.0 | 3.7 | 5.8 |
| In US <15 year | 6.1 | 1.4 | 5.3 | 18.9 | 31.1 | 2.6 | 1.1 |
| DK | 0.3 | 0.1 | 0.3 | 1.0 | 0.9 | 0.1 | 0.2 |
| Language of interview |  |  |  |  |  |  |  |
| English | 93.8 | 99.7 | 99.7 | 64.0 | 92.4 | 99.4 | 99.9 |
| Other | 6.2 | 0.4 | 0.3 | 36.0 | 7.6 | 0.6 | 0.1 |
| Marital status |  |  |  |  |  |  |  |
| Married | 53.4 | 56.9 | 33.7 | 51.6 | 64.8 | 40.0 | 36.9 |
| Previously married | 19.6 | 20.4 | 23.8 | 15.7 | 10.8 | 24.0 | 20.4 |
| Never married | 26.8 | 22.6 | 42.3 | 32.5 | 24.3 | 35.8 | 42.5 |
| DK | 0.2 | 0.2 | 0.2 | 0.2 | 0.2 | 0.2 | 0.1 |
| Children at home |  |  |  |  |  |  |  |
| No children | 61.0 | 65.2 | 59.9 | 46.7 | 52.4 | 56.8 | 67.3 |
| One child | 17.2 | 16.1 | 19.2 | 19.3 | 21.2 | 18.6 | 14.9 |
| Two or more | 21.8 | 18.7 | 20.9 | 34.0 | 26.4 | 24.6 | 17.9 |
| Education |  |  |  |  |  |  |  |
| Less than HS or GED | 15.6 | 10.9 | 18.6 | 35.4 | 9.2 | 25.2 | 14.1 |
| HS diploma | 22.4 | 22.3 | 26.0 | 22.8 | 14.6 | 23.0 | 20.4 |
| Some college | 19.7 | 20.0 | 23.1 | 17.5 | 13.7 | 23.8 | 28.2 |
| Associate degree | 11.2 | 12.0 | 10.8 | 9.1 | 7.5 | 11.3 | 11.7 |
| Bachelor's degree | 19.6 | 22.0 | 14.0 | 10.1 | 31.3 | 10.5 | 16.2 |
| Master's or more | 11.2 | 12.6 | 7.0 | 4.2 | 23.2 | 5.1 | 9.3 |
| DK | 0.4 | 0.3 | 0.5 | 0.8 | 0.6 | 1.0 | 0.2 |
| Family income |  |  |  |  |  |  |  |
| \$0-34,999 | 28.5 | 23.9 | 42.6 | 38.4 | 23.0 | 45.6 | 35.7 |
| \$35,000-74,999 | 28.5 | 28.3 | 28.2 | 31.7 | 23.4 | 26.8 | 26.6 |
| \$75,000-99,999 | 11.6 | 12.7 | 8.5 | 9.3 | 11.8 | 9.1 | 11.1 |
| \$100,000+ | 23.7 | 27.6 | 12.6 | 13.1 | 33.0 | 11.2 | 20.9 |
| DK | 7.6 | 7.5 | 8.1 | 7.6 | 8.9 | 7.4 | 5.8 |
| Home ownership |  |  |  |  |  |  |  |
| Owns home | 66.0 | 73.8 | 47.2 | 49.8 | 60.8 | 55.9 | 54.5 |
| Rents | 31.7 | 23.9 | 49.9 | 48.0 | 37.0 | 41.0 | 41.9 |
| Other | 2.4 | 2.3 | 2.9 | 2.2 | 2.2 | 3.1 | 3.6 |
| N | 273,972 | 172,782 | 36,357 | 43,160 | 15,289 | 1,920 | 4,464 |

Weighted percent shown for all variables except age where weighted mean is shown.
Design-adjusted F-tests show that all variables differ significantly across the race/ethnic groups at $p < .001$ .
All racial categories other than Hispanic are Non-Hispanic. AIAN=American Indian/Alaska Native, or Native American.

Next, we examine whether and how the pain disparities are influenced by the different distributions of covariates across race/ethnicity, by estimating a series of five logistic regression models for each of the six pain measures (Table 4). We control for only year of interview in Model 1, add age, age squared, gender, and region of residence in Model 2, nativity and language in Model 3, marital status and children in Model 4, and, finally, we add the three socioeconomic measures in Model 5. For parsimony, Table 4 shows only the odds ratios associated with race/ethnicity in the document below; the full models with all coefficients are in Supplemental Table S4.

**Table 4.** Odds ratios from logistic models of six pain indicators as a function of race and covariates

| White=reference | Model 1 | Model 2 | Model 3 | Model 4 | Model 5 |
| --- | --- | --- | --- | --- | --- |
| Severe pain |  |  |  |  |  |
| Black | 1.13* | 1.21** | 1.25*** | 1.14* | 0.84** |
| Hispanic | 0.72*** | 0.88* | 1.12 | 1.08 | 0.80* |
| Asian | 0.34*** | 0.38*** | 0.54*** | 0.55*** | 0.60*** |
| AIAN | 1.71** | 1.90*** | 1.90*** | 1.79** | 1.15 |
| Multiracial | 1.31* | 1.70*** | 1.70*** | 1.58*** | 1.31* |
| High-impact pain |  |  |  |  |  |
| Black | 0.95 | 1.08 | 1.12 | 1.01 | 0.74*** |
| Hispanic | 0.74*** | 0.95 | 1.18 | 1.13 | 0.85 |
| Asian | 0.40*** | 0.45*** | 0.67** | 0.68** | 0.71* |
| AIAN | 1.93*** | 2.17*** | 2.20*** | 1.97*** | 1.30 |
| Multiracial | 1.51*** | 2.02*** | 2.05*** | 1.93*** | 1.56*** |
| Widespread pain |  |  |  |  |  |
| Black | 0.82*** | 0.84*** | 0.86*** | 0.81*** | 0.66*** |
| Hispanic | 0.75*** | 0.79*** | 0.97 | 0.96 | 0.80*** |
| Asian | 0.38*** | 0.37*** | 0.51*** | 0.52*** | 0.56*** |
| AIAN | 1.42*** | 1.37*** | 1.37*** | 1.32** | 1.00 |
| Multiracial | 1.54*** | 1.69*** | 1.70*** | 1.64*** | 1.45*** |
| Persistent pain (3 months) |  |  |  |  |  |
| Black | 0.71*** | 0.79*** | 0.82*** | 0.77*** | 0.63*** |
| Hispanic | 0.51*** | 0.62*** | 0.81*** | 0.79*** | 0.65*** |
| Asian | 0.32*** | 0.35*** | 0.51*** | 0.51*** | 0.54*** |
| AIAN | 1.41** | 1.61*** | 1.62*** | 1.57*** | 1.17 |
| Multiracial | 1.00 | 1.32** | 1.33** | 1.28** | 1.14 |
| Persistent pain (6 months) |  |  |  |  |  |
| Black | 0.72*** | 0.80*** | 0.83*** | 0.79*** | 0.63*** |
| Hispanic | 0.56*** | 0.69*** | 0.89 | 0.86* | 0.70*** |
| Asian | 0.28*** | 0.31*** | 0.45*** | 0.46*** | 0.48*** |
| AIAN | 1.08 | 1.16 | 1.18 | 1.11 | 0.82 |
| Multiracial | 1.14 | 1.45*** | 1.48*** | 1.43*** | 1.24* |
| Any pain |  |  |  |  |  |
| Black | 0.74*** | 0.80*** | 0.82*** | 0.81*** | 0.72*** |
| Hispanic | 0.62*** | 0.70*** | 0.89*** | 0.88*** | 0.80*** |
| Asian | 0.45*** | 0.46*** | 0.61*** | 0.61*** | 0.64*** |
| AIAN | 1.00 | 1.02 | 1.03 | 1.01 | 0.88 |
| Multiracial | 1.10* | 1.28*** | 1.30*** | 1.28*** | 1.20*** |

\*\*\*p<.001 \*\*p<.01 \*p<.05
The table shows partial results from survey-adjusted logistic regression model of each pain outcome as a function of race/ethnicity (White as reference), net of five sets of covariates. Only the odds ratios associated with race/ethnicity are shown for parsimony; the complete results are in the supplement.
All racial categories other than Hispanic are Non-Hispanic. AIAN=American Indian/Alaska Native, or Native American.
Model 1 only adjusts for the year of interview.
Model 2 additionally adjusts for age, gender, and region of residence.
Model 3 additionally adjusts for immigrant status and language of interview.
Model 4 additionally adjusts for marital status and presence of children.
Model 5 is fully adjusted; it additionally adjusts for education, family income, and home ownership.

These models frame the disparities in relative terms, using odds ratios. To visualize the findings and show the disparities in absolute terms, we also calculate average predicted probabilities of each pain level from the demographics-adjusted Model 2; the estimates and their 95% confidence intervals are in Figure 1. We also visualize the predicted probabilities from fully adjusted models – that is, net of all demographic and socioeconomic characteristics – and show the resulting set of plots in Supplement Figure 1.

**Figure 1.**
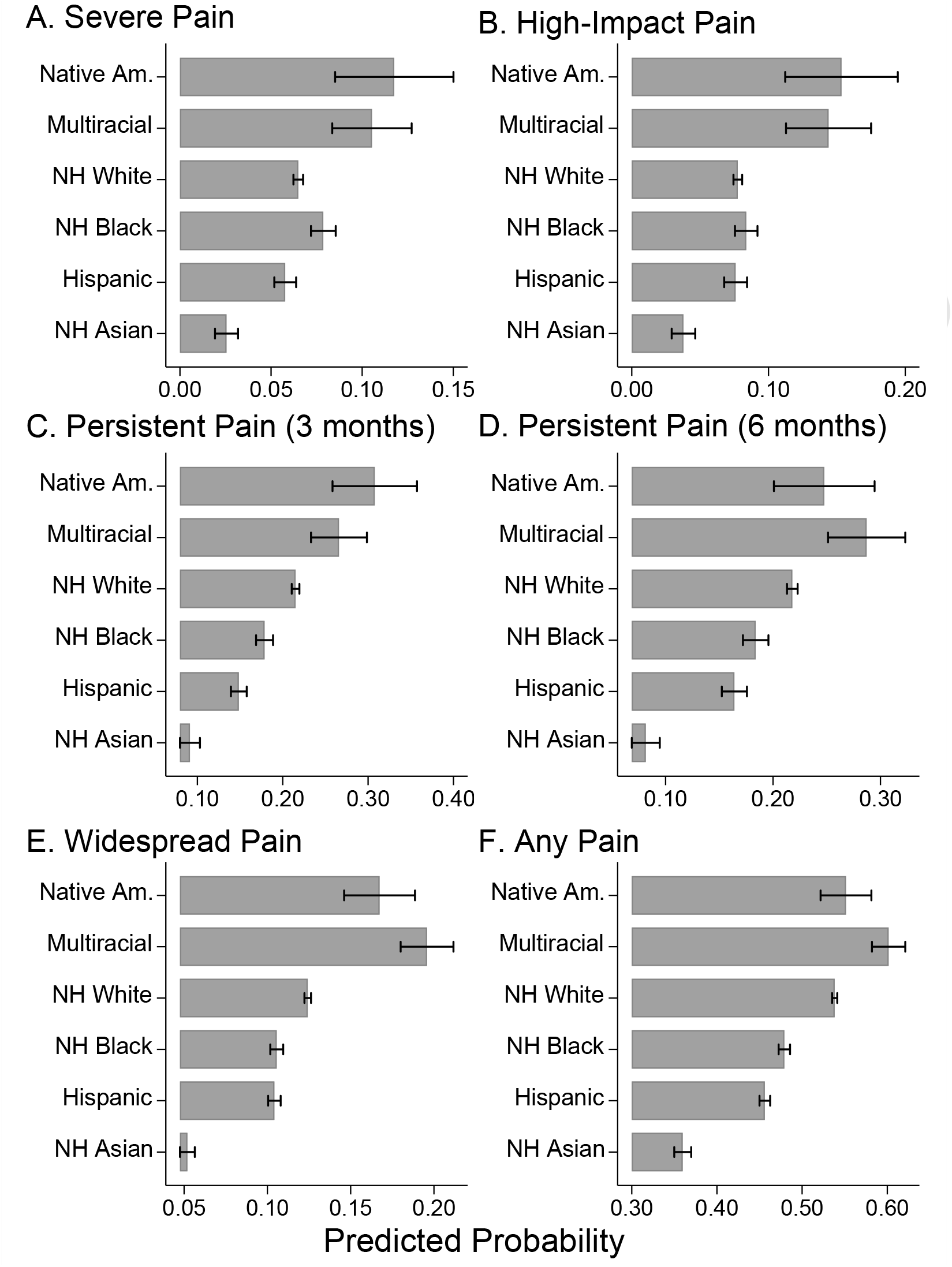
Average predicted probabilities, from age, sex, and year-adjusted models (Model 2 Table 4) Average predicted probabilities calculated from complex-survey-adjusted logistic models of each pain measure as a function of race/ethnicity, year of interview, age (centered about 60) and age squared, gender, and region (Model 2 in Table 4)

Finally, we attend to the fact that the analyses above include all ages 18 and older. Age has a major impact on pain levels [16], as well as the distribution of most pain covariates [55]. However, the models above force the effect of race/ethnicity to be equal across the full adult age range. As a final step, we therefore examine how the racial/ethnic patterns in each pain measure differ by age. To do so in a flexible, data-driven way, we use the plreg command in Stata [27] to estimate race/ethnicity-stratified semiparametric partial-linear models of the form *P*_*i*_ = *α* + *f*(*a*_*i*_) + *γx*_*i*_, where *P*_*i*_ is the presence of pain (yes=1 versus no=0); *x*_*i*_ is a vector of covariates (we control for year and gender); and *a*_*i*_ captures age. The smooth function of age *f*(*a*_*i*_) is estimated by the lowess procedure in Stata [9]. This model thus estimates the level of pain across age for each racial/ethnic group, while additively including other covariates. The results are plotted as line graphs of pain across age in Figure 2.

**Figure 2.**
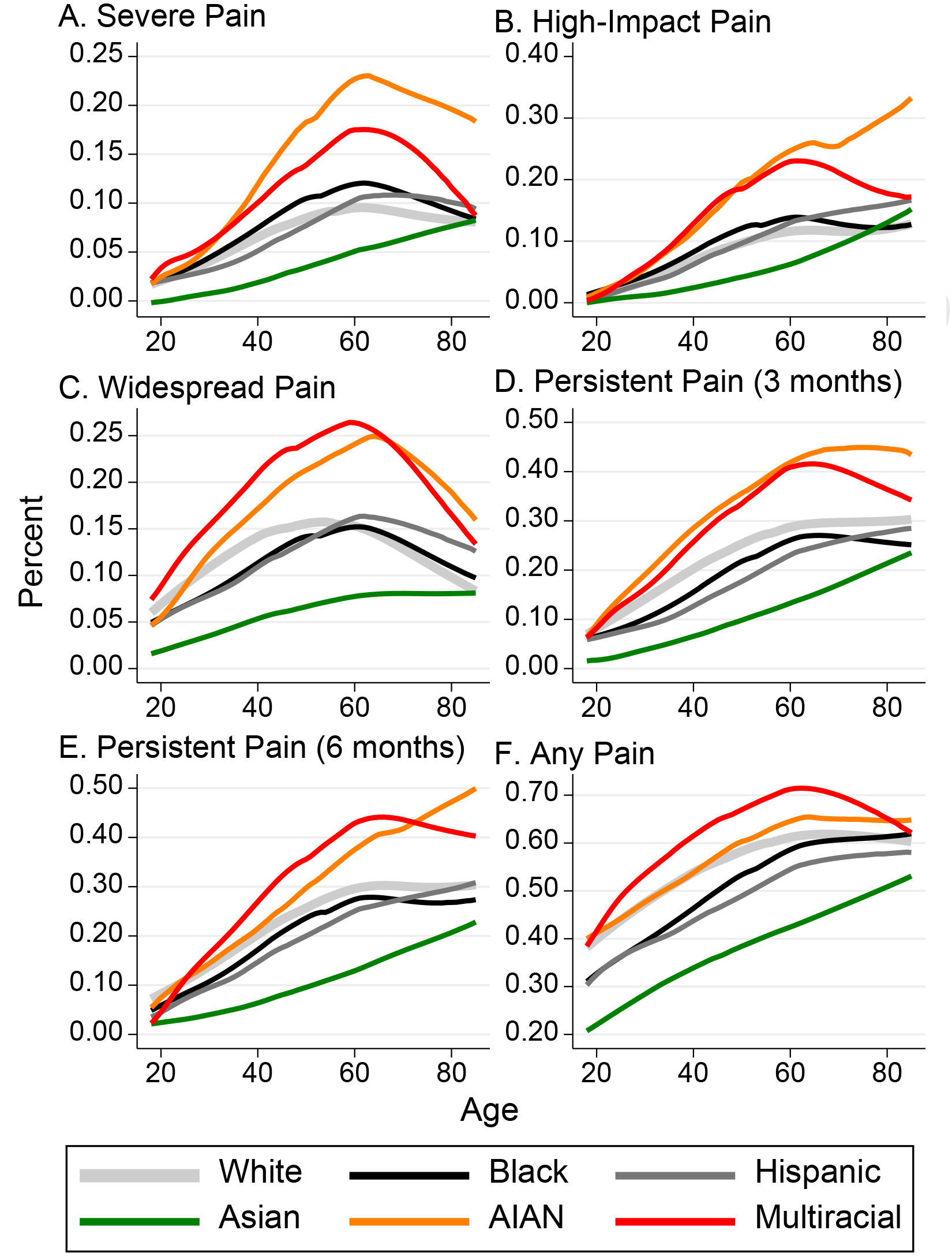
Estimated pain levels by age for each race/ethnic group. Note: figure based on semiparametric models of each pain definition, net of year of interview and gender and a flexibly estimated age function

The analytic steps outlined above thus summarize the gross pain disparities by race/ethnicity and show the disparities net of important potential confounders. The findings are presented in relative terms, as odds ratios comparing each non-White group vis-à-vis Whites, and in absolute results, as predicted probabilities of pain in each group. Jointly, the analysis yields a comprehensive portrait of the national racial distribution of chronic pain.

## RESULTS

Table 2 shows the weighted prevalence of pain in the total sample as well as in the six racial/ethnic groups for all six definitions of pain. The prevalence for each pain indicator is color-coded for easier interpretation, ranging from green (for the group with the lowest prevalence for a given pain measure) to red (highest prevalence). Additionally, the six pain measures are ordered, left to right, from the lowest to the highest total prevalence.

Several findings regarding the estimated prevalence in the total adult population stand out. First, the prevalence ranges widely, from 6.4% (95%CI 6.2,6.7) for severe pain to 51.0% (95%CI 50.7, 51.4) for ‘any pain.’ As well, high-impact pain affects 7.7% of the population (95%CI 7.4, 8.0), while widespread pain and persistent pain are experienced by about 12% and 20% of the American population, respectively. Second, the prevalence of persistent pain is effectively identical for a 3-month versus 6-month horizon (19.6% versus 20.0%). This finding is consistent with prior reports, in which 3-versus 6-month time horizons “appear to have little or no effect on prevalence estimates” [45] perhaps because of imprecise recall of the onset and duration of the pain [3; 37]. This finding also suggests that estimates from either question may be treated as comparable, which may be important for pain scholars who only have access to one of these measures in population surveys.

With respect to racial/ethnic disparities, the estimates are mostly consistent across all pain measures. By far the lowest prevalence of each pain indicator is among Asian Americans. The two groups with the highest prevalence, in contrast, are Native American and multiracial adults. White, Black, and Hispanic groups fall in between these two extremes. The disparities are tremendous. Severe pain is reported by 2.4% of Asian Americans (95%CI 1.8,3.0), 5.0% of Hispanics (95%CI 4.5,5.5), 6.8% of Whites (95%CI 6.5,7.1), 7.6% of Blacks (95%CI 6.9,8.3), 8.7% of multiracial adults (95%CI 6.9,10.6), and 11.1% of Native Americans (95%CI 7.5,14.7). In other words, Hispanics have double the prevalence of severe pain compared to Asian Americans, multiracial adults have 3.6 times the prevalence, and Native Americans have an astonishing 4.6 times the prevalence.

Moreover, high-impact chronic pain occurs in 3.5% of Asian Americans, but 14.7% of Native Americans (4.2 times higher) and 12.0% of multiracial adults (3.5 times higher). White and Black adults have about 8% probability (8.3% and 7.9% respectively) of high-impact pain, while Hispanic adults have a probability of 6.3%. Similar patterns are evident for the less stringent pain operationalization, although the differences become smaller in relative terms and larger in absolute terms. For instance, multiracial adults have 1.6 times the prevalence of ‘any pain’ compared with Asian Americans, but the difference is almost 22 percentage points higher (57.1%) among multiracial Americans versus 35.3% among Asian Americans.

The final note from Table 2 is that the relative ‘position’ of White adults changes from the most to the least impactful pain operationalizations: for the most severe definitions, Whites have less or equal prevalence of pain than Black adults and only modestly more than Hispanic adults. For the least restrictive definition (any pain), Whites have substantially higher pain not only relative to Black and Hispanic groups, but equal to that of Native Americans. The shading highlights this pattern: as the pain definitions progress from the most to the least stringent, the shading indicates increasingly higher prevalence among Whites relative the total population.

Supplemental Table S1 shows that these general patterns hold for both genders: Asian men and women have the lowest pain prevalence, while Native American and multiracial men and women have the highest. The results also corroborate the well-established pattern of higher pain among women compared to men [4]. In all but three of 42 available comparisons across groups and pain definitions, women had higher pain prevalence than men, and in many cases, the difference was large. For instance, the prevalence of severe pain was 5.3% among men (95%CI 5.0,5.6) but 42% higher, or 7.5% (95%CI 7.4,8.2), among women. The largest relative excess pain was for widespread pain, which is about 33% to 88% greater among women across all racial/ethnic categories, compared with their male counterparts. Finally, we note that the intersection of gender and race/ethnicity produces enormous disparities. For instance, severe pain is experienced by only 2.2% of Asian American men, but 12.9% of Native American women (5.9 times the prevalence), and high-impact pain is experienced by 3.2% of Asian American men but 12.8% of both Native American and multiracial women (4 times the prevalence).

Supplemental Table S2 adds nuance to the racial/ethnic comparisons by disaggregating the multiracial category by the main racial background reported by multiracial respondents Overall, there was statistically significant heterogeneity across these subgroups for severe, widespread, and both persistent pain measures, but not for high-impact pain or ‘any pain.’ The most noteworthy finding is that multiracial adults who listed Native American as their main racial background had by far the highest pain prevalence of any group for all pain measures: 13.2% reported severe pain (95%CI 7.2,19.3), 22.8% reported high-impact chronic pain (95%CI 11.6,33.9), and 31.6% reported widespread pain (95%CI 25.5,37.6), in comparison to 6.4%, 7.7%, and 11.6% in the total U.S. population, respectively. The multiracial adults who reported their main racial group as White also had relatively high pain across a number of pain measures, while the pain prevalence of other groups varied. A part of this variability may be due to the relatively modest sample sizes (although Supplemental Table S3 shows that the number of respondents in each subcategory exceeded 100 for all but multiracial Asian adults for high-impact and persistent pain over 6 months), but we believe that the findings should spur additional research into pain among multiracial adults.

Table 3 summarizes the characteristics of the target population across race and ethnicity. This table is relevant because some of the racial/ethnic group differences described above may be attributed to different sociodemographic characteristics if their distribution varies across groups. Indeed, we found that every covariate differed statistically significantly across the groups (p<.001). For instance, Whites were older (mean age 49.0 years) than all other groups, while Hispanic (40.8) and multiracial (40.4) adults were the youngest. For the other two basic demographic factors, sex differences were more modest across groups but region of residence differed substantially. Another significant area of difference was immigrant status, where well over 90% of White, Native American, and multiracial adults were U.S.-born, while only 42.9% of Hispanic and 22.1% of Asian Americans were. Correspondingly, nearly all White, Native American, and multiracial adults conducted the interview in English, compared with only 64% of Hispanics. Among Asian Americans, despite the high proportion of immigrants in their population (especially those with less than 15 years in the U.S.), nearly 93% completed the interview in English. Finally, socioeconomic status (SES), whether captured with education, family income, or home ownership, differed markedly as well. In terms of education and family income, Asian Americans had the highest rates, while Hispanic and Native Americans had the lowest. For home ownership, Whites had the highest rates, while Black Americans had the lowest.

The next step, therefore, was to adjust the gross comparisons for these sociodemographic differences. Table 4 summarizes the findings from a series of logistic regression models for each of the six pain measures, gradually adjusted for all characteristics. For parsimony, the table only shows odds ratios for each racial/ethnic group compared to the White reference group, across all models and pain measures. The full results showing the effects of all covariates are in Supplemental Table S4.

Asian Americans’ have significantly lower pain vis-à-vis the reference category of Whites across all pain outcomes and net of all covariate sets (Models 1 through 5). Net of age, gender, and region of residence, they have 62% lower odds of severe pain, 55% lower odds of high-impact pain, and 63% lower odds of widespread pain, compared to Whites. Controlling for Asian American’s high proportion of immigrants, as well as any differences in terms of family composition and socioeconomic status in Models 3-5, the differences attenuate but remain statistically significant and substantively large for all pain measures.

On the other end of the pain spectrum are Native American and multiracial adults. Native Americans have roughly twice the odds of severe and high-impact pain compared to Whites, net of demographics (Model 2, OR=1.90, p<.001, for severe pain and OR=2.17, p<.001, for high-impact pain). The pain ‘excess’ is also evident and statistically significant for widespread pain and persistent pain over 3 months, although not for persistent pain over 6 months and ‘any pain.’ Nativity and family composition do not change these patterns meaningfully. Importantly, however, when we control for socioeconomic status in Model 5, the excess pain becomes not significant. That is, the significantly higher pain in Native Americans, relative to Whites, is explained by their lower SES.

Further, the excess pain among multiracial adults is significant in 27 out of 30 models across all six pain definitions and most covariate sets. Multiracial adults have 70% higher odds of severe pain, and 100% higher odds of high-impact pain, compared to Whites, net of year, age, gender, and region (Model 2). Unlike for Native Americans, moreover, this excess remains statistically significant and substantively large even net of SES, although about half the excess is explained by the covariates. For instance, even in the fully adjusted Model 5, multiracial Americans have 31% higher odds of severe pain, 56% higher odds of high-impact pain, and 45% higher odds of widespread pain, compared to Whites.

Hispanic Americans have either significantly lower pain than Whites (for most pain measures and across most models) or the difference between the groups is not statistically significant. The Hispanics pain ‘advantage’ is smallest in Model 3 where we control for immigrant status and language of interview. Family composition had only a modest effect but controlling for the Hispanics’ lower SES, vis-à-vis Whites, enlarged the Hispanic advantage. That is, Hispanics, when compared to Whites, report significantly less pain, suggesting this advantage stems from factors other than those we included in the analysis.

Finally, the comparison between Black and White adults differs between pain measures. The most stringent pain measure, severe pain, shows Black adults have significantly more pain compared to Whites (OR=1.21, p<.01, Model 2). However, this excess of severe pain becomes significantly lower if we take into account the Black-White SES differences. That is, net of education, family income, and home ownership in Model 5, Black Americans have significantly *less* severe pain (OR=0.84, p<.01) than Whites. There is no significant difference between Black and White adults for the second most stringent pain measure, high-impact chronic pain, until we control for SES (net of SES differences, Blacks have significantly less pain, OR=0.74, p<.001). For the remaining four less stringent pain operationalizations, Black adults have statistically significantly less pain than Whites, no matter what characteristics we control for across the models.

In addition to the racial/ethnic differences, the models yield several important results with respect to the covariates (shown in Supplement Table S4). In particular, they corroborate well-recognized patterns, wherein pain increases with age, but at a decreasing rate (that is, there may be a plateau or even decline in pain at older ages in these cross-sectional models; although it is important to remember this pattern may be a function of selective mortality of cohort differences rather than true declines with age [7; 16]); women report significantly more pain than men; and immigrants report significantly less pain than native-born persons. The models also show the powerful association between socioeconomic status and pain: for all pain measures, each of the three SES measures (education, family income, and home ownership) were significant correlates of pain in the expected direction.

We visualized the average predicted probabilities of each pain measure across the race/ethnic groups in Figure 1 and Supplemental Figure S1. Figure 1 shows the probability of pain net of year, age, gender, and region of residence (Model 2); Supplemental Figure S1 shows the probability of pain net of all covariates (Model 5). The figures show the differences across the groups in absolute terms, highlighting the high pain prevalence among Native American and multiracial adults, and the low pain prevalence among Asian Americans.

Finally, we examined the pain prevalence in each racial/ethnic group across age, using a semiparametric approach with a flexible function for age and visualizing the resulting age curves in Figure 2. The main finding is that the results described above are generally valid across the adult life span. That is, there is little evidence of systematic ‘crossovers,’ whereby the overall results would only be valid for some but not other parts of the life span. The differences across race/ethnic groups are relatively small at age 18 because pain prevalence is relatively low for all groups in early adulthood. The prevalence, as well as racial/ethnic differences, increase until early older ages (roughly age 60), when at least some groups and at least some pain measures show a plateau or a decline in prevalence with advanced ages. These results corroborate prior research with cross-sectional data where, as a function of mortality selection and/or rising pain across successive birth cohorts, pain prevalence may appear to decrease at older ages [8; 16]. Interestingly, Asian Americans show no evidence of a plateau or even decelerating increase with age: pain prevalence increases almost linearly across age through to 84. In contrast, multiracial adults have the most pronounced apex around age 60 and lower pain at ages beyond that threshold. Other groups are in between, evidencing some combination of steady increases, plateauing, or decreases in pain at the oldest ages.

## DISCUSSION

Our analysis provides the first comprehensive portrait of pain prevalence across all major U.S. racial/ethnic groups, including the fastest-growing multiracial category, for an array of six pain measures ranging from ‘severe’ and ‘high-impact’ pain to the broadly defined ‘any pain.’ The findings build on prior work on racial/ethnic differences in pain, especially among Black and White adults, and provide new understanding of on pain among Hispanic, Asian American, Native American, and multiracial groups.

Asian Americans have the lowest pain prevalence of all groups, with a statistically significant and substantial pain ‘advantage.’ That is, they have less than half the probability of pain compared with the U.S. average for all pain measures except ‘any pain,’ where they still have 30% lower prevalence. This pattern aligns with the few prior population studies that included Asian American and other groups [30; 58]. Additionally, our analysis shows that Asian Americans’ low prevalence is observed across all pain severity levels, and persists net of demographic and socioeconomic differences.

In contrast, Native American (AIAN) and multiracial adults have higher pain levels than all other groups. Native Americans have by far the highest prevalence of severe pain and high-impact pain, 11.1% and 14.7%, respectively, versus overall U.S. averages of 6.4% and 7.7%. These estimates substantially expand the limited body of scholarship documenting Native Americans’ high pain levels [12; 21; 39; 43]. Compared to Whites, this pain prevalence remains significantly higher, even when we control for age, sex, immigrant status, language, and family covariates. However, when we statistically equalize socioeconomic status differences, the pain disadvantage becomes not significant. That is, the excess pain among Native Americans can be attributed to their disadvantaged socioeconomic position vis-à-vis Whites. As Meghani [29] cautions, this result does not mean that there is no racial disparity, but rather that the societal conditions that produce the pain disparities are inextricably grounded in the “lack of equality of social opportunities” (p. 2224).

Americans self-identifying as multiracial also suffer disproportionately from pain. They have the highest or second-highest prevalence on all pain measures. Unlike among Native Americans, however, controlling for SES attenuates but does not explain this excess pain among multiracial adults. That is, multiracial Americans experience significantly more pain than Whites, and the causes lie beyond lower SES. Moreover, we discovered a racial ‘intersectionality’ effect [35]: the intersection of being multiracial and considering Native American as one‘s main racial background is associated with the highest pain prevalence of any group by far. Little is known about multiracial individuals’ health: research is limited and focuses on adolescents’ mental health and health behaviors, not on adults’ health, let alone pain [5]. Our troubling findings should serve as impetus for focused examination of pain in this rapidly growing U.S. group.

White, Black, and Hispanic adults fall between these two extremes. Of these three groups, Hispanics have the lowest pain prevalence for all pain measures -- significantly less than, or comparable to, Whites. This echoes some prior work describing lower pain among Hispanics than Whites [18; 22; 30; 55] but contradicts other research that found higher pain in this group [16; 38]. The majority of Hispanic American adults are foreign-born (57%). The socially disadvantaged status of immigrants, some have theorized, predisposes them to increased pain [10]. However, empirical work has shown that U.S. immigrants have *less* pain than the U.S.-born [17; 55], a pattern our analyses corroborated. Correspondingly, controlling for immigrant status attenuated the pain ‘advantage’ for Hispanics. However, it did not fully ‘explain’ the lower pain prevalence of Hispanics relative to Whites. That is, only a minor part of the Hispanic pain advantage appears linked to the high proportion of immigrants; much of the difference remains unexplained by the factors considered in our study (the same reasoning and empirical findings are also evident for Asian Americans).

Finally, the comparison between Black and White Americans is particularly important for two reasons. First, their comparison dominates scholarship on racial/ethnic pain disparities, yet these the pain differences between these two groups are modest relative to disparities across other groups. It is unclear why Black Americans, who have a much higher prevalence of physical health problems than Whites [52], don‘t have correspondingly higher levels of pain as well; this question should be addressed in future research (perhaps a clue lies in Blacks’ lower rates of *mental* health problems [28]). Second, the Black-White differences are complex, in that they depend on the pain definition. For the least stringent ‘any pain’ and persistent pain measures, Blacks have significantly less pain than Whites, about 26-29% lower odds. For high-impact pain, Blacks and Whites have statistically the same prevalence, and for the most stringent pain definition of severe pain, Black adults have significantly *higher* prevalence than Whites (25% higher odds, net of demographics). This pattern echoes prior work that reported higher prevalence of severe pain for Blacks but lower prevalence for mild pain [11; 33; 56]. Our set of results clarifies that such seemingly inconsistent findings may simply reflect differences in pain operationalizations. Reasons for this pattern are unclear. Perhaps Blacks are less likely than Whites to report milder pain, but more likely to report severe pain. However, an equally valid interpretation is that Whites are particularly likely to experience and report some pain, including mild pain, but less likely to experience more severe pain. Indeed, this interpretation is better supported by our findings, which show Whites increasingly likely to report pain vis-à-vis all Americans for milder pain definitions. One additional point about the Black-White disparities is the critical role of SES. If we compare adults at the same SES levels, Blacks have less pain than Whites across all six pain measures, even severe pain. This finding highlights the inextricable role of social factors in pain and pain disparities; however, it also leaves open the question of why Blacks at comparable levels of SES report systematically lower pain than Whites. This is another important direction for further research.

Of note, our findings regarding population pain prevalence are somewhat at odds with the literature on experimental pain, which often finds Black, Asian, and Hispanic participants to have greater pain sensitivity than Whites [2; 14; 24; 32], while the prevalence of naturally occurring pain in the population is lower. Paradoxically, Native Americans do not seem to have higher pain sensitivity than whites [40], yet they have higher prevalence, as we just described. This pattern of results suggests that findings from experimental pain research may not provide accurate indications of pain disparities in the general population.

We point out several limitations of our study. Although the NHIS includes a rich set of variables, the data nonetheless omit important information. For instance, NHIS lacks detailed information about the combined racial/ethnic self-identification of multiracial adults. Others have found that specific combinations of race/ethnicity matter for pain and health in general [5; 31]; such information would allow us to better distinguish which racial/ethnic combinations (not only which primary race) are associated with vulnerability to pain. Furthermore, important groups such as Arab Americans are not categorized in NHIS, although over 2 million Americans are of Arab ancestry [49]. This group has experienced sharply increased discrimination and stigmatization since 9/11 [1], which may increase susceptibility to chronic pain [10]. Moreover, the NHIS data lacks information about perceived discrimination and other race-related potential stressors. We theorize that these characteristics could be important risk factors for pain in racial minority adults. For instance, multiracial Americans are subject to unique forms of discrimination, such as that based on a racial categorization that does not match the individual’s self-identity [15] – a type of discrimination associated with particularly poor health outcomes [51]. Future studies should collect data on perceived discrimination that could shed light on these processes.

The biopsychosocial model of pain predicts that that people “marginalized by social conditions” would have more pain [10]. Our foundational results show more nuanced patterns, in which some minoritized groups have higher pain prevalence than Whites, while others have lower prevalence. We hope these findings motivate further in-depth studies of drivers of racial/ethnic differences in pain risk, to inform policy, prevention, and intervention efforts. Indeed, given that pain is arguably the most prevalent and costly public health condition in the U.S., enhanced knowledge of racial and ethnic disparities in pain is urgently needed to inform policy decisions and focus efforts at population-level prevention and intervention. Pain epidemiology must become more comprehensive—moving beyond a focus on Black-White comparisons—to characterize and understand pain disparities in the increasingly diverse U.S. population.

## Data Availability

The data in this analysis are all publicly available at https://nhis.ipums.org/nhis/

## ACKNOWLEDGEMENTS

We thank Dr. Richard Nahin, Dr. Zachary Zimmer, Dr. Jason Winders, Dr. Autumn Knowlton, Avery Smith, and Lindsay Finlay for their assistance, suggestions, and comments on various drafts of this manuscript.

Research reported in this publication was supported by the National Institute on Aging of the National Institutes of Health under Award Number R01AG065351 (PI: Grol-Prokopczyk), and by the Social Sciences and Humanities Research Council of Canada (SSHRC) Insight Grant (PI: Zajacova). The content is solely the responsibility of the authors and does not necessarily represent the official views of the National Institutes of Health or SSHRC.

All authors declare no conflict of interest.

## SUPPLEMENT

### Sample sizes

We analyze six pain measures. The information was collected in different years and on different subsamples; the sample sizes differ accordingly, from N=57,518 for high-impact chronic pain to 273,972 for ‘any pain’ and widespread pain. Table 1 in the main document describes the sample size, years of collection, and construction of each of the six pain measures.

This paragraph provides additional details on the sample sizes for each of the six measures. Any pain and widespread pain items were collected from all sample adults in each year 2010-2018 (274,868 respondents). From this group, 99.7% answered all five pain questions needed for the two summary pain indicators (any pain and widespread pain, see below) and also provided valid information on race. This yields <u>N=273,972</u> used in all analyses of any pain and widespread pain (383 individuals or 0.01% had one or more components missing on pain while an additional 512 individuals or 0.02% did not answer the race question). Second, persistent pain over 6 months and high-impact chronic pain were collected in a special module administered in 2016 and 2017 to 57,737 adults. Of this group, 99.7% answered both pain frequency and race items, yielding an analytic sample of size <u>N=57,541</u> for persistent pain over 6 months (64 respondents or 0.01% did not answer the pain frequency question and 132 or 0.02% did not provide a valid race information). An additional 23 individuals (<.01%) did not answer a followup questions about interference, yielding a sample size of <u>N=57,518</u> (99.6% of those administered the items) for analyses of high-impact chronic pain. Third, data on persistent pain over 3 months and severe pain were collected consistently in years 2010-2015 and 2018 (104,156 respondents were administered the questions). From this group, 99.3% provided valid information on the pain frequency and also race, yielding a sample size of <u>N=103,439</u> for analyses of persistent pain over 3 months (521 or 0.5% did not answer the pain frequency question, and additional 186 did not provide valid race answer). Finally, an additional 90 (0.09%) respondents did not answer a followup question on pain intensity, which was necessary to construct the severe pain measure, yielding a sample size of <u>N=103,349</u> for analyses of severe pain, or 99.2% of the targeted sample.

**Supplement Table S1.**
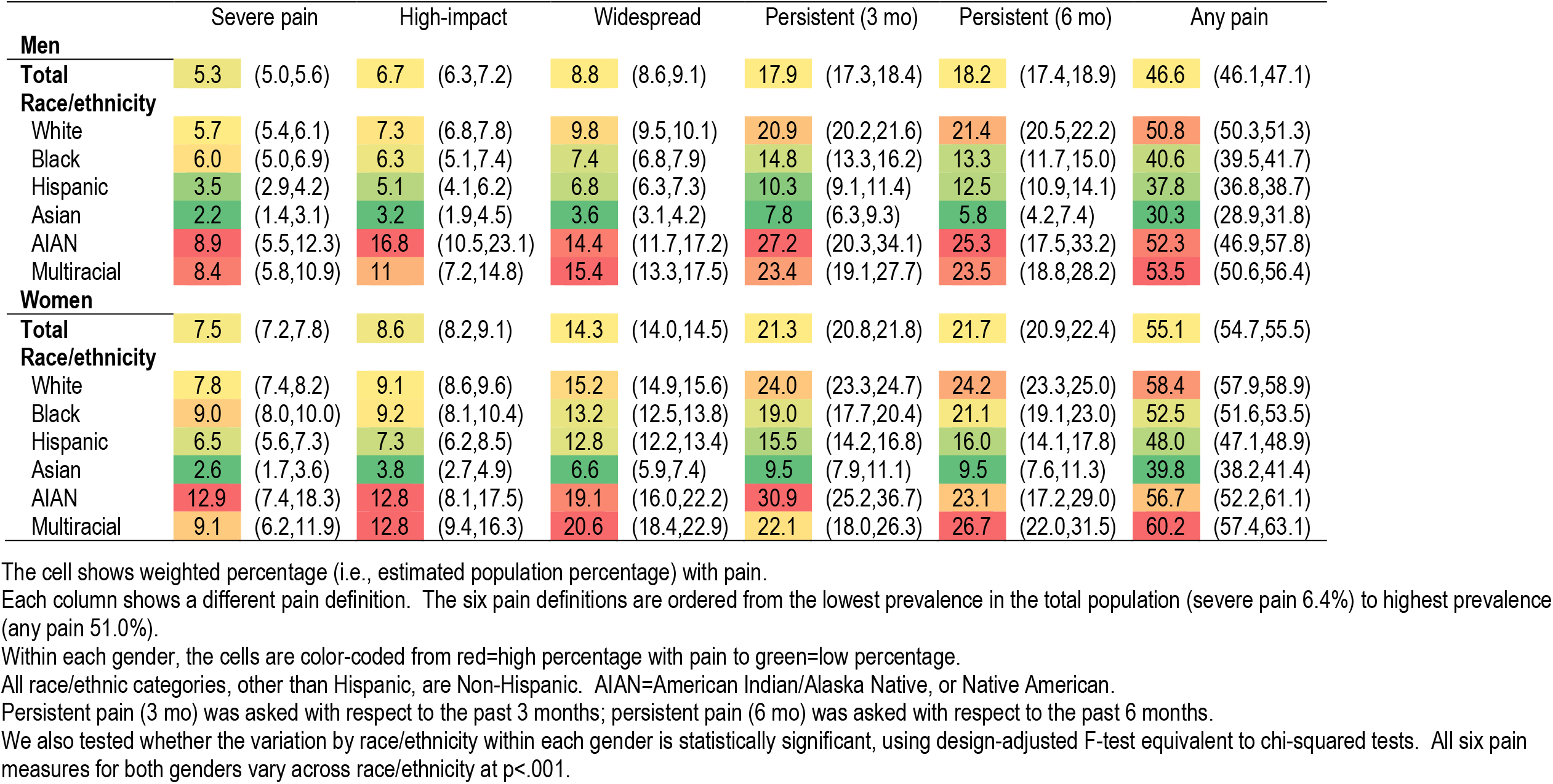
Weighted proportion with pain across different pain measures and associated sample sizes by gender, US adults age 18+.

**Supplement Table S2.** Weighted percent with pain (95% CI) across different pain measures, multiracial US adults age 18+ by main stated racial background.

|  | Severe pain |  | High-impact |  | Widespread |  | Persistent (3 mo) |  | Persistent (6 mo) |  | Any pain |  |
| --- | --- | --- | --- | --- | --- | --- | --- | --- | --- | --- | --- | --- |
| **Total** | 6.4 | (6.2,6.7) | 7.7 | (7.4,8.0) | 11.6 | (11.4,11.8) | 19.6 | (19.2,20.0) | 20.0 | (19.4,20.6) | 51.0 | (50.7,51.4) |
| **Multiracial adults, by main racial background** |  |  |  |  |  |  |  |  |  |  |  |  |
| White | 11.4 | (8.4,14.5) | 11.2 | (8.0,14.5) | 20.7 | (18.4,23.1) | 26.5 | (21.9,31.1) | 28.0 | (22.9,33.0) | 61.2 | (58.0,64.3) |
| Black | 7.4 | (3.2,11.7) | 8.4 | (3.9,12.8) | 13.8 | (10.9,16.7) | 18.2 | (11.9,24.5) | 17.1 | (11.0,23.3) | 51.0 | (46.5,55.5) |
| Asian | 0.0 | (0.0,0.0) | 19.6 | (-0.6,39.8) | 7.5 | (4.1,10.8) | 14.0 | (1.7,26.3) | 26.9 | (7.4,46.4) | 49.3 | (41.1,57.6) |
| AIAN | 13.2 | (7.2,19.3) | 22.8 | (11.6,33.9) | 31.6 | (25.5,37.6) | 29.5 | (19.8,39.3) | 38.8 | (25.6,52.0) | 68.7 | (62.7,74.7) |
| Other | 3.8 | (1.4,6.2) | 9.9 | (3.9,15.8) | 13.2 | (10.3,16.2) | 18.1 | (11.6,24.6) | 20.2 | (13.3,27.2) | 50.1 | (45.2,55.0) |
| **All racial/ethnic groups for comparison, estimates identical to those shown in Table 2** |  |  |  |  |  |  |  |  |  |  |  |  |
| White | 6.8 | (6.5,7.1) | 8.3 | (7.9,8.6) | 12.6 | (12.4,12.8) | 22.5 | (22.0,23.0) | 22.8 | (22.1,23.5) | 54.7 | (54.3,55.1) |
| Black | 7.6 | (6.9,8.3) | 7.9 | (7.1,8.7) | 10.6 | (10.1,11.0) | 17.1 | (16.1,18.1) | 17.6 | (16.2,19.0) | 47.2 | (46.4,48.0) |
| Hispanic | 5.0 | (4.5,5.5) | 6.3 | (5.4,7.1) | 9.8 | (9.4,10.2) | 12.9 | (12.0,13.7) | 14.2 | (12.9,15.5) | 42.9 | (42.1,43.6) |
| Asian | 2.4 | (1.8,3.0) | 3.5 | (2.6,4.4) | 5.2 | (4.8,5.7) | 8.7 | (7.6,9.7) | 7.7 | (6.6,8.9) | 35.3 | (34.2,36.5) |
| AIAN | 11.1 | (7.5,14.7) | 14.7 | (10.5,18.9) | 17.0 | (14.8,19.3) | 29.3 | (24.7,33.9) | 24.2 | (18.7,29.6) | 54.7 | (50.9,58.5) |
| Multiracial | 8.7 | (6.9,10.6) | 12.0 | (9.4,14.5) | 18.2 | (16.6,19.7) | 22.7 | (19.8,25.7) | 25.2 | (21.9,28.4) | 57.1 | (54.9,59.2) |

**Supplement Table S3.** Sample sizes for all analyses, US adults age 18+.

|  | Severe pain | High-impact pain | Widespread pain | Persistent pain (3 mo) | Persistent pain (6 mo) | Any pain |
| --- | --- | --- | --- | --- | --- | --- |
| Total | 103,349 | 57,518 | 273,972 | 103,439 | 57,541 | 273,972 |
| Race/ethnicity |  |  |  |  |  |  |
| White | 64,629 | 40,324 | 172,782 | 64,694 | 40,342 | 172,782 |
| Black | 13,749 | 6,086 | 36,357 | 13,759 | 6,089 | 36,357 |
| Hispanic | 16,698 | 6,800 | 43,160 | 16,709 | 6,801 | 43,160 |
| Asian | 5,816 | 2,815 | 15,289 | 5,818 | 2,815 | 15,289 |
| AIAN | 743 | 482 | 1,920 | 744 | 483 | 1,920 |
| Multiracial | 1,714 | 1,011 | 4,464 | 1,715 | 1,011 | 4,464 |
| Multiracial |  |  |  |  |  |  |
| White | 766 | 477 | 1,977 | 767 | 477 | 1,977 |
| Black | 375 | 163 | 911 | 375 | 163 | 911 |
| Asian | 106 | 68 | 303 | 106 | 68 | 303 |
| AIAN | 164 | 107 | 455 | 164 | 107 | 455 |
| Other | 303 | 196 | 818 | 303 | 196 | 818 |

All categories other than Hispanic are Non-Hispanic. AIAN=American Indian/Alaska Native, or Native American. For parsimony, we omit sample sizes for men and women; each gender comprises roughly 50% of the total or each race/ethnic group.
All race/ethnic categories, other than Hispanic, are Non-Hispanic. AIAN=American Indian/Alaska Native, or Native American.
Persistent pain (3 mo) was asked with respect to the past 3 months; persistent pain (6 mo) was asked with respect to the past 6 months.

**Supplement Table S4.**
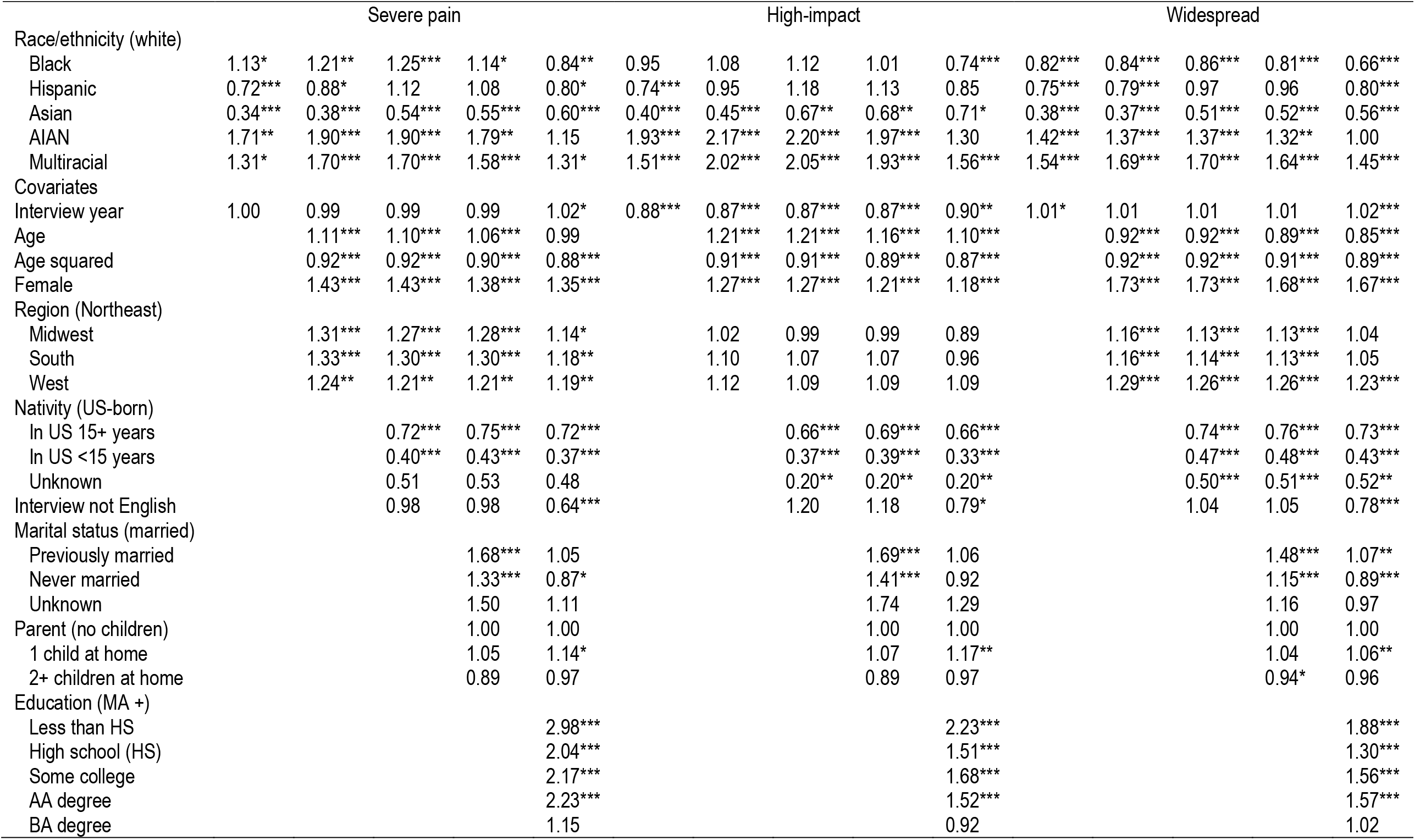
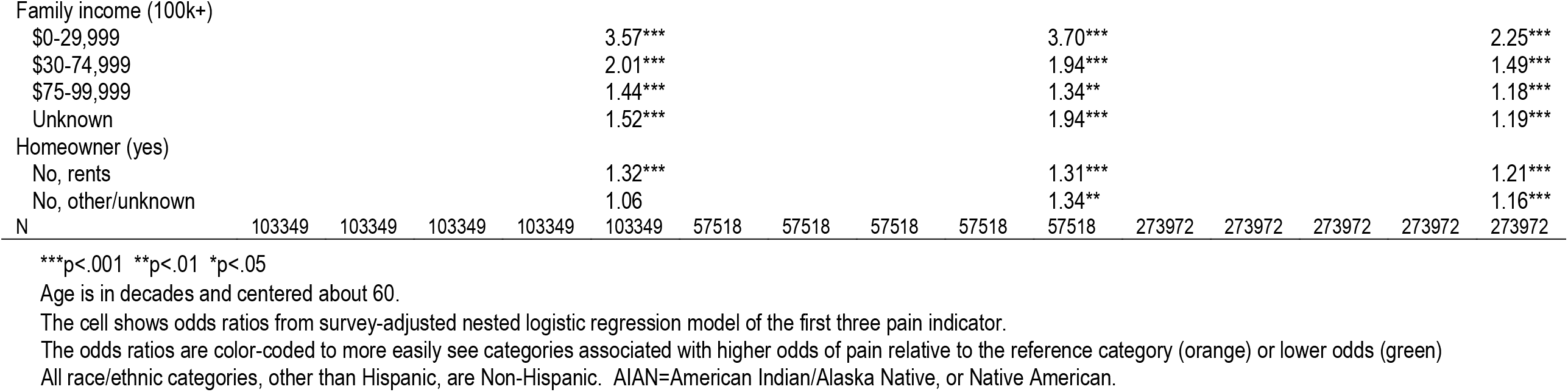
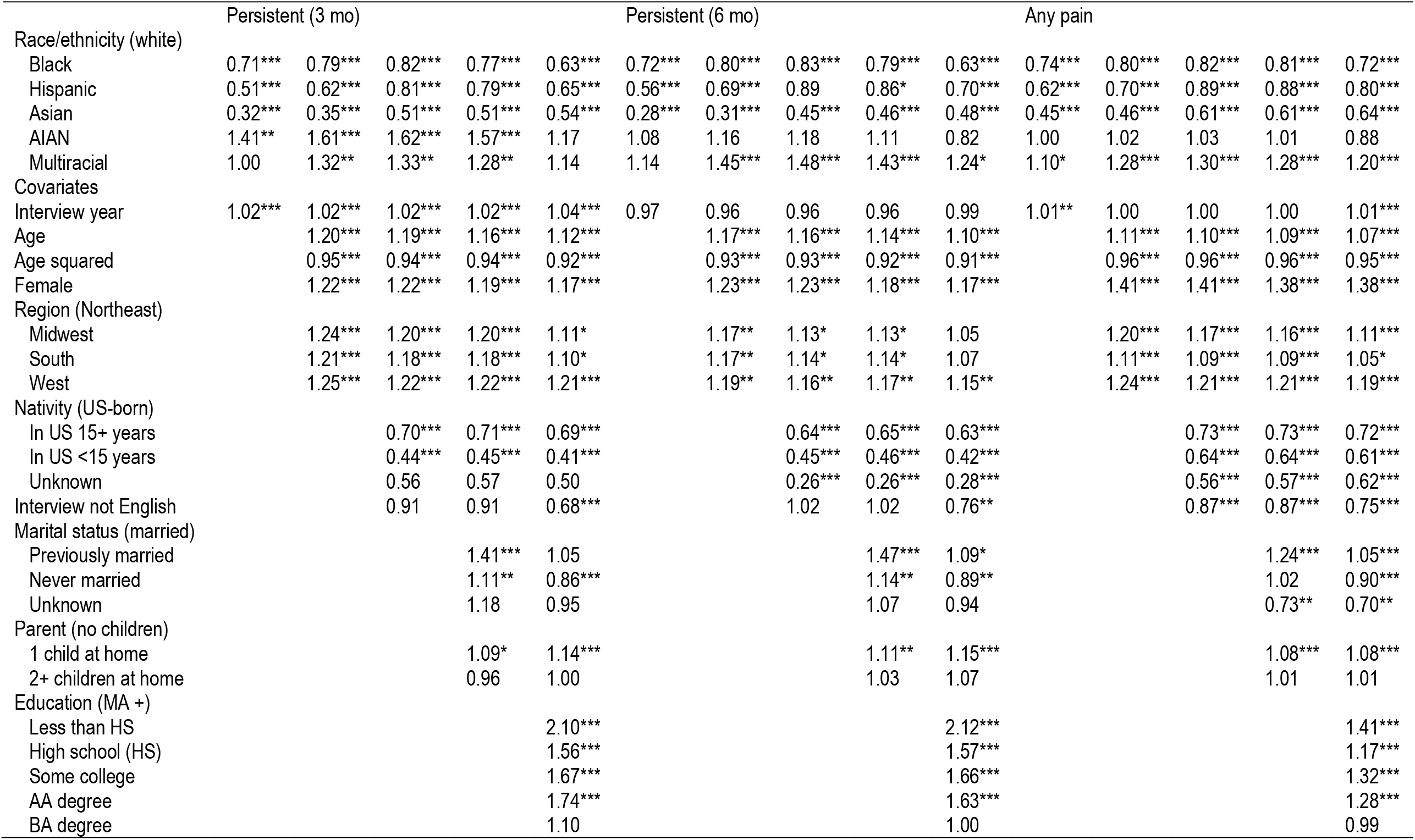
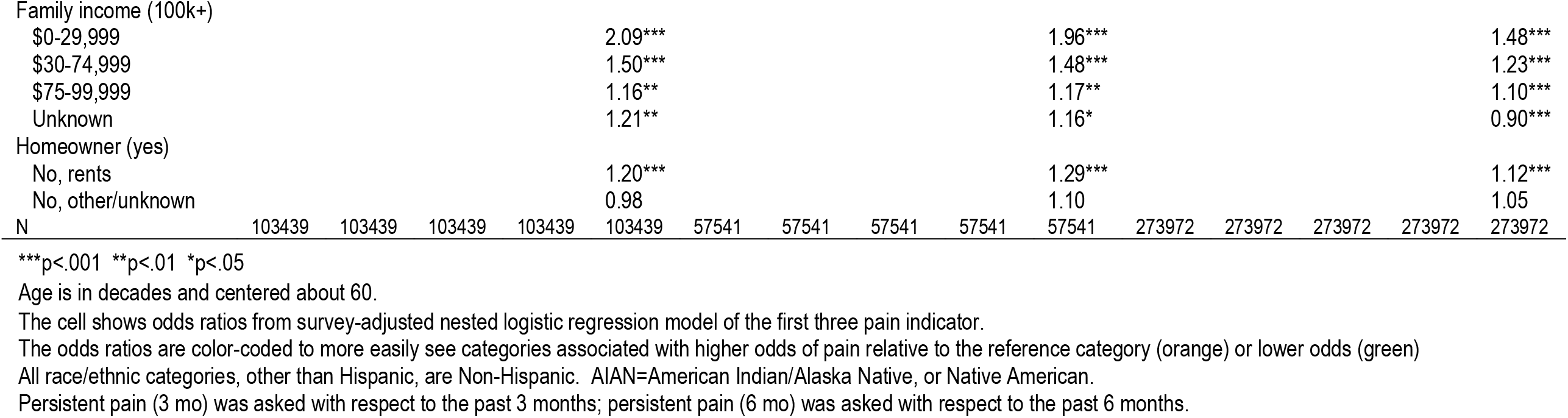
Odds ratios from logistic regression models of each pain measure as a function of race/ethnicity and different covariate sets

|  | Severe pain |  |  |  |  | High-impact |  |  |  |  | Widespread |  |  |  |  |
| --- | --- | --- | --- | --- | --- | --- | --- | --- | --- | --- | --- | --- | --- | --- | --- |
| Race/ethnicity (white) |  |  |  |  |  |  |  |  |  |  |  |  |  |  |  |
| Black | 1.13* | 1.21** | 1.25*** | 1.14* | 0.84** | 0.95 | 1.08 | 1.12 | 1.01 | 0.74*** | 0.82*** | 0.84*** | 0.86*** | 0.81*** | 0.66*** |
| Hispanic | 0.72*** | 0.88* | 1.12 | 1.08 | 0.80* | 0.74*** | 0.95 | 1.18 | 1.13 | 0.85 | 0.75*** | 0.79*** | 0.97 | 0.96 | 0.80*** |
| Asian | 0.34*** | 0.38*** | 0.54*** | 0.55*** | 0.60*** | 0.40*** | 0.45*** | 0.67** | 0.68** | 0.71* | 0.38*** | 0.37*** | 0.51*** | 0.52*** | 0.56*** |
| AIAN | 1.71** | 1.90*** | 1.90*** | 1.79** | 1.15 | 1.93*** | 2.17*** | 2.20*** | 1.97*** | 1.30 | 1.42*** | 1.37*** | 1.37*** | 1.32** | 1.00 |
| Multiracial | 1.31* | 1.70*** | 1.70*** | 1.58*** | 1.31* | 1.51*** | 2.02*** | 2.05*** | 1.93*** | 1.56*** | 1.54*** | 1.69*** | 1.70*** | 1.64*** | 1.45*** |
| Covariates |  |  |  |  |  |  |  |  |  |  |  |  |  |  |  |
| Interview year | 1.00 | 0.99 | 0.99 | 0.99 | 1.02* | 0.88*** | 0.87*** | 0.87*** | 0.87*** | 0.90** | 1.01* | 1.01 | 1.01 | 1.01 | 1.02*** |
| Age |  | 1.11*** | 1.10*** | 1.06*** | 0.99 |  | 1.21*** | 1.21*** | 1.16*** | 1.10*** |  | 0.92*** | 0.92*** | 0.89*** | 0.85*** |
| Age squared |  | 0.92*** | 0.92*** | 0.90*** | 0.88*** |  | 0.91*** | 0.91*** | 0.89*** | 0.87*** |  | 0.92*** | 0.92*** | 0.91*** | 0.89*** |
| Female |  | 1.43*** | 1.43*** | 1.38*** | 1.35*** |  | 1.27*** | 1.27*** | 1.21*** | 1.18*** |  | 1.73*** | 1.73*** | 1.68*** | 1.67*** |
| Region (Northeast) |  |  |  |  |  |  |  |  |  |  |  |  |  |  |  |
| Midwest |  | 1.31*** | 1.27*** | 1.28*** | 1.14* |  | 1.02 | 0.99 | 0.99 | 0.89 |  | 1.16*** | 1.13*** | 1.13*** | 1.04 |
| South |  | 1.33*** | 1.30*** | 1.30*** | 1.18** |  | 1.10 | 1.07 | 1.07 | 0.96 |  | 1.16*** | 1.14*** | 1.13*** | 1.05 |
| West |  | 1.24** | 1.21** | 1.21** | 1.19** |  | 1.12 | 1.09 | 1.09 | 1.09 |  | 1.29*** | 1.26*** | 1.26*** | 1.23*** |
| Nativity (US-born) |  |  |  |  |  |  |  |  |  |  |  |  |  |  |  |
| In US 15+ years |  |  | 0.72*** | 0.75*** | 0.72*** |  |  | 0.66*** | 0.69*** | 0.66*** |  |  | 0.74*** | 0.76*** | 0.73*** |
| In US <15 years |  |  | 0.40*** | 0.43*** | 0.37*** |  |  | 0.37*** | 0.39*** | 0.33*** |  |  | 0.47*** | 0.48*** | 0.43*** |
| Unknown |  |  | 0.51 | 0.53 | 0.48 |  |  | 0.20** | 0.20** | 0.20** |  |  | 0.50*** | 0.51*** | 0.52** |
| Interview not English |  |  | 0.98 | 0.98 | 0.64*** |  |  | 1.20 | 1.18 | 0.79* |  |  | 1.04 | 1.05 | 0.78*** |
| Marital status (married) |  |  |  |  |  |  |  |  |  |  |  |  |  |  |  |
| Previously married |  |  |  | 1.68*** | 1.05 |  |  |  | 1.69*** | 1.06 |  |  |  | 1.48*** | 1.07** |
| Never married |  |  |  | 1.33*** | 0.87* |  |  |  | 1.41*** | 0.92 |  |  |  | 1.15*** | 0.89*** |
| Unknown |  |  |  | 1.50 | 1.11 |  |  |  | 1.74 | 1.29 |  |  |  | 1.16 | 0.97 |
| Parent (no children) |  |  |  | 1.00 | 1.00 |  |  |  | 1.00 | 1.00 |  |  |  | 1.00 | 1.00 |
| 1 child at home |  |  |  | 1.05 | 1.14* |  |  |  | 1.07 | 1.17** |  |  |  | 1.04 | 1.06** |
| 2+ children at home |  |  |  | 0.89 | 0.97 |  |  |  | 0.89 | 0.97 |  |  |  | 0.94* | 0.96 |
| Education (MA +) |  |  |  |  |  |  |  |  |  |  |  |  |  |  |  |
| Less than HS |  |  |  |  | 2.98*** |  |  |  |  | 2.23*** |  |  |  |  | 1.88*** |
| High school (HS) |  |  |  |  | 2.04*** |  |  |  |  | 1.51*** |  |  |  |  | 1.30*** |
| Some college |  |  |  |  | 2.17*** |  |  |  |  | 1.68*** |  |  |  |  | 1.56*** |
| AA degree |  |  |  |  | 2.23*** |  |  |  |  | 1.52*** |  |  |  |  | 1.57*** |
| BA degree |  |  |  |  | 1.15 |  |  |  |  | 0.92 |  |  |  |  | 1.02 |
| Family income (100k+) |  |  |  |  |  |  |  |  |  |  |  |  |  |  |  |
| \$0-29,999 | | | | | 3.57*** | | | | | 3.70*** | | | | | 2.25*** |
| \$30-74,999 | | | | | 2.01*** | | | | | 1.94*** | | | | | 1.49*** |
| \$75-99,999 | | | | | 1.44*** | | | | | 1.34** | | | | | 1.18*** |
| Unknown |  |  |  |  | 1.52*** |  |  |  |  | 1.94*** |  |  |  |  | 1.19*** |
| Homeowner (yes) |  |  |  |  |  |  |  |  |  |  |  |  |  |  |  |
| No, rents |  |  |  |  | 1.32*** |  |  |  |  | 1.31*** |  |  |  |  | 1.21*** |
| No, other/unknown |  |  |  |  | 1.06 |  |  |  |  | 1.34** |  |  |  |  | 1.16*** |
| N | 103349 | 103349 | 103349 | 103349 | 103349 | 57518 | 57518 | 57518 | 57518 | 57518 | 273972 | 273972 | 273972 | 273972 | 273972 |

\*\*\*p<.001 \*\*p<.01 \*p<.05

| Continued Supplement Table S4. Odds ratios from logistic regression models of each pain measure as a function of race/ethnicity and different covariate sets |  |  |  |  |  |  |  |  |  |  |  |  |  |  |  |
| --- | --- | --- | --- | --- | --- | --- | --- | --- | --- | --- | --- | --- | --- | --- | --- |
|  | Persistent (3 mo) |  |  |  |  | Persistent (6 mo) |  |  |  |  | Any pain |  |  |  |  |
| Race/ethnicity (white) |  |  |  |  |  |  |  |  |  |  |  |  |  |  |  |
| Black | 0.71*** | 0.79*** | 0.82*** | 0.77*** | 0.63*** | 0.72*** | 0.80*** | 0.83*** | 0.79*** | 0.63*** | 0.74*** | 0.80*** | 0.82*** | 0.81*** | 0.72*** |
| Hispanic | 0.51*** | 0.62*** | 0.81*** | 0.79*** | 0.65*** | 0.56*** | 0.69*** | 0.89 | 0.86* | 0.70*** | 0.62*** | 0.70*** | 0.89*** | 0.88*** | 0.80*** |
| Asian | 0.32*** | 0.35*** | 0.51*** | 0.51*** | 0.54*** | 0.28*** | 0.31*** | 0.45*** | 0.46*** | 0.48*** | 0.45*** | 0.46*** | 0.61*** | 0.61*** | 0.64*** |
| AIAN | 1.41** | 1.61*** | 1.62*** | 1.57*** | 1.17 | 1.08 | 1.16 | 1.18 | 1.11 | 0.82 | 1.00 | 1.02 | 1.03 | 1.01 | 0.88 |
| Multiracial | 1.00 | 1.32** | 1.33** | 1.28** | 1.14 | 1.14 | 1.45*** | 1.48*** | 1.43*** | 1.24* | 1.10* | 1.28*** | 1.30*** | 1.28*** | 1.20*** |
| Covariates |  |  |  |  |  |  |  |  |  |  |  |  |  |  |  |
| Interview year | 1.02*** | 1.02*** | 1.02*** | 1.02*** | 1.04*** | 0.97 | 0.96 | 0.96 | 0.96 | 0.99 | 1.01** | 1.00 | 1.00 | 1.00 | 1.01*** |
| Age |  | 1.20*** | 1.19*** | 1.16*** | 1.12*** |  | 1.17*** | 1.16*** | 1.14*** | 1.10*** |  | 1.11*** | 1.10*** | 1.09*** | 1.07*** |
| Age squared |  | 0.95*** | 0.94*** | 0.94*** | 0.92*** |  | 0.93*** | 0.93*** | 0.92*** | 0.91*** |  | 0.96*** | 0.96*** | 0.96*** | 0.95*** |
| Female |  | 1.22*** | 1.22*** | 1.19*** | 1.17*** |  | 1.23*** | 1.23*** | 1.18*** | 1.17*** |  | 1.41*** | 1.41*** | 1.38*** | 1.38*** |
| Region (Northeast) |  |  |  |  |  |  |  |  |  |  |  |  |  |  |  |
| Midwest |  | 1.24*** | 1.20*** | 1.20*** | 1.11* |  | 1.17** | 1.13* | 1.13* | 1.05 |  | 1.20*** | 1.17*** | 1.16*** | 1.11*** |
| South |  | 1.21*** | 1.18*** | 1.18*** | 1.10* |  | 1.17** | 1.14* | 1.14* | 1.07 |  | 1.11*** | 1.09*** | 1.09*** | 1.05* |
| West |  | 1.25*** | 1.22*** | 1.22*** | 1.21*** |  | 1.19** | 1.16** | 1.17** | 1.15** |  | 1.24*** | 1.21*** | 1.21*** | 1.19*** |
| Nativity (US-born) |  |  |  |  |  |  |  |  |  |  |  |  |  |  |  |
| In US 15+ years |  |  | 0.70*** | 0.71*** | 0.69*** |  |  | 0.64*** | 0.65*** | 0.63*** |  |  | 0.73*** | 0.73*** | 0.72*** |
| In US <15 years |  |  | 0.44*** | 0.45*** | 0.41*** |  |  | 0.45*** | 0.46*** | 0.42*** |  |  | 0.64*** | 0.64*** | 0.61*** |
| Unknown |  |  | 0.56 | 0.57 | 0.50 |  |  | 0.26*** | 0.26*** | 0.28*** |  |  | 0.56*** | 0.57*** | 0.62*** |
| Interview not English |  |  | 0.91 | 0.91 | 0.68*** |  |  | 1.02 | 1.02 | 0.76** |  |  | 0.87*** | 0.87*** | 0.75*** |
| Marital status (married) |  |  |  |  |  |  |  |  |  |  |  |  |  |  |  |
| Previously married |  |  |  | 1.41*** | 1.05 |  |  |  | 1.47*** | 1.09* |  |  |  | 1.24*** | 1.05*** |
| Never married |  |  |  | 1.11** | 0.86*** |  |  |  | 1.14** | 0.89** |  |  |  | 1.02 | 0.90*** |
| Unknown |  |  |  | 1.18 | 0.95 |  |  |  | 1.07 | 0.94 |  |  |  | 0.73** | 0.70** |
| Parent (no children) |  |  |  |  |  |  |  |  |  |  |  |  |  |  |  |
| 1 child at home |  |  |  | 1.09* | 1.14*** |  |  |  | 1.11** | 1.15*** |  |  |  | 1.08*** | 1.08*** |
| 2+ children at home |  |  |  | 0.96 | 1.00 |  |  |  | 1.03 | 1.07 |  |  |  | 1.01 | 1.01 |
| Education (MA +) |  |  |  |  |  |  |  |  |  |  |  |  |  |  |  |
| Less than HS |  |  |  |  | 2.10*** |  |  |  |  | 2.12*** |  |  |  |  | 1.41*** |
| High school (HS) |  |  |  |  | 1.56*** |  |  |  |  | 1.57*** |  |  |  |  | 1.17*** |
| Some college |  |  |  |  | 1.67*** |  |  |  |  | 1.66*** |  |  |  |  | 1.32*** |
| AA degree |  |  |  |  | 1.74*** |  |  |  |  | 1.63*** |  |  |  |  | 1.28*** |
| BA degree |  |  |  |  | 1.10 |  |  |  |  | 1.00 |  |  |  |  | 0.99 |
| Family income (100k+) |  |  |  |  |  |  |  |  |  |  |  |  |  |  |  |
| \$0-29,999 | | | | | 2.09*** | | | | | 1.96*** | | | | | 1.48*** |
| \$30-74,999 | | | | | 1.50*** | | | | | 1.48*** | | | | | 1.23*** |
| \$75-99,999 | | | | | 1.16** | | | | | 1.17** | | | | | 1.10*** |
| Unknown |  |  |  |  | 1.21** |  |  |  |  | 1.16* |  |  |  |  | 0.90*** |
| Homeowner (yes) |  |  |  |  |  |  |  |  |  |  |  |  |  |  |  |
| No, rents |  |  |  |  | 1.20*** |  |  |  |  | 1.29*** |  |  |  |  | 1.12*** |
| No, other/unknown |  |  |  |  | 0.98 |  |  |  |  | 1.10 |  |  |  |  | 1.05 |
| N | 103439 | 103439 | 103439 | 103439 | 103439 | 57541 | 57541 | 57541 | 57541 | 57541 | 273972 | 273972 | 273972 | 273972 | 273972 |

\*\*\*p<.001 \*\*p<.01 \*p<.05
Age is in decades and centered about 60.
The cell shows odds ratios from survey-adjusted nested logistic regression model of the first three pain indicator.
The odds ratios are color-coded to more easily see categories associated with higher odds of pain relative to the reference category (orange) or lower odds (green)
All race/ethnic categories, other than Hispanic, are Non-Hispanic. AIAN=American Indian/Alaska Native, or Native American.
Persistent pain (3 mo) was asked with respect to the past 3 months; persistent pain (6 mo) was asked with respect to the past 6 months.

**Supplement Figure S1.**
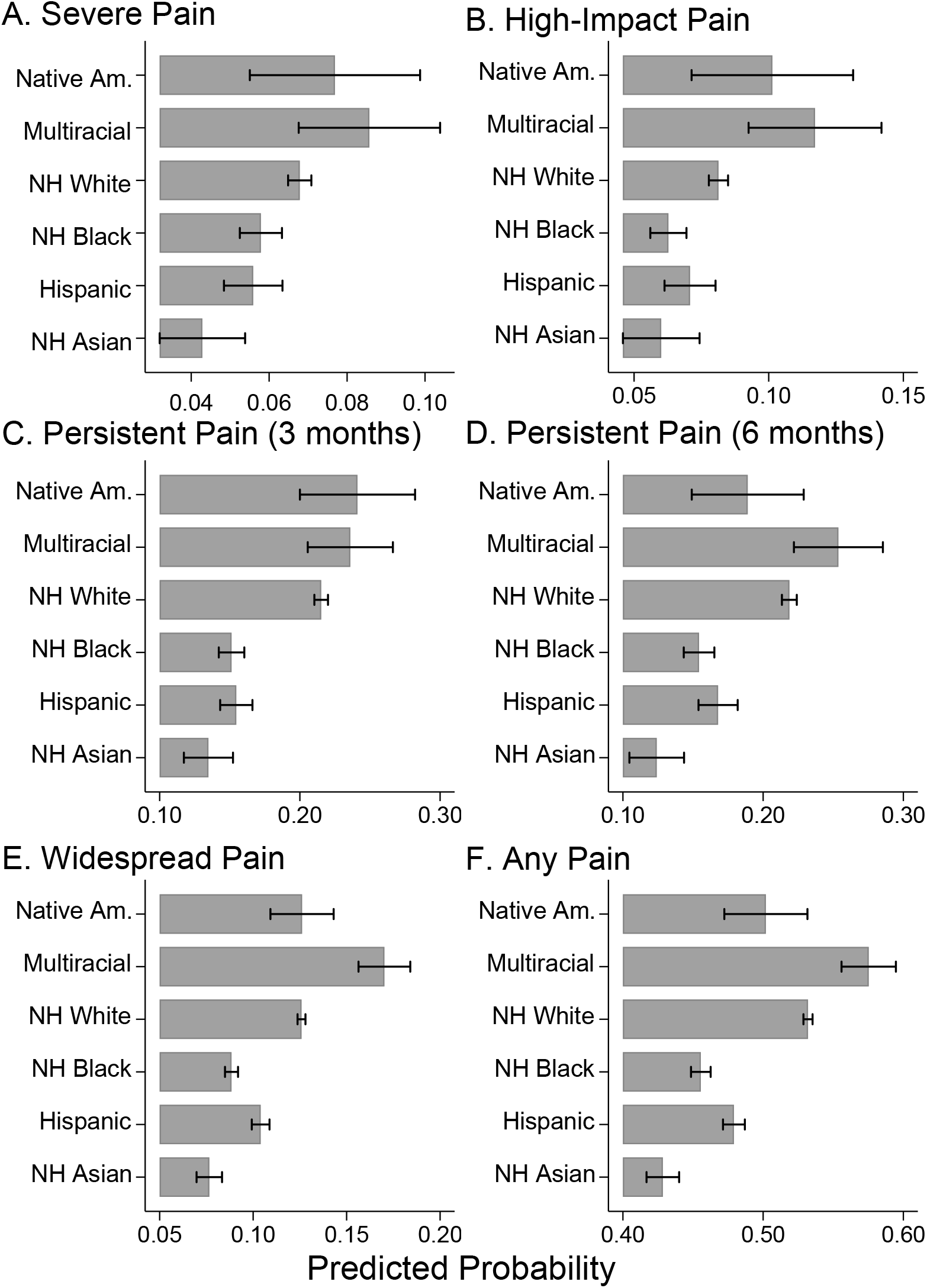
Average predicted probabilities from fully adjusted models. Average predicted probabilities calculated from complex-survey-adjusted logistic models of each pain measure as a function of race/ethnicity and all covariates (Model 5 Table 4)

## REFERENCES

[1] Abuelezam NN, El-Sayed AM, Galea S. The Health of Arab Americans in the United States: An Updated Comprehensive Literature Review. Frontiers in Public Health 2018;6(262):1–18.

[2] Ahn H, Weaver M, Lyon DE, Kim J, Choi E, Staud R, Fillingim RB. Differences in Clinical Pain and Experimental Pain Sensitivity Between Asian Americans and Whites With Knee Osteoarthritis. The Clinical Journal of Pain 2017;33(2).

[3] Barsky AJ. Forgetting, Fabricating, and Telescoping: The Instability of the Medical History. Archives of Internal Medicine 2002;162(9):981–984.

[4] Bartley EJ, Fillingim RB. Sex differences in pain: a brief review of clinical and experimental findings. British Journal of Anaesthesia 2013;111(1):52–58.

[5] Bratter JL, Mason C. Multiracial Americans, Health Patterns, and Health Policy: Assessment and Recommendations for Ways Forward. In: KO Korgen, editor. Race Policy and Multiracial Americans. Bristol, UK: Policy Press, 2016. pp. 155–171.

[6] Campbell CM, Edwards RR. Ethnic differences in pain and pain management. Pain Management 2012;2(3):219–230.

[7] Case A, Deaton A. The Misery and Mystery of Pain. Deaths of Despair and the Future of Capitalism. Princeton NJ: Princeton University Press, 2020. pp. 83–93.

[8] Case A, Deaton A, Stone AA. Decoding the mystery of American pain reveals a warning for the future. Proceedings of the National Academy of Sciences 2020;117(40):24785–24789.

[9] Cleveland WS. Robust Locally Weighted Regression and Smoothing Scatterplots. Journal of the American Statistical Association 1979;74(368):829–836.

[10] Craig KD, Holmes C, Hudspith M, Moor G, Moosa-Mitha M, Varcoe C, Wallace B. Pain in persons who are marginalized by social conditions. PAIN(r) 2020;161(2):261–265.

[11] Dahlhamer J, Lucas J, Zelaya C, Nahin R, Mackey S, DeBar L, Kerns R, Von Korff M, Porter L, Helmick C. Prevalence of chronic pain and high-impact chronic pain among adults—United States, 2016. Morbidity and Mortality Weekly Report 2018;67(36):1–6.

[12] Deyo RA, Mirza SK, Martin BI. Back pain prevalence and visit rates: estimates from U.S. national surveys, 2002. Spine 2006;31(23):2724–2727.

[13] Edwards CL, Fillingim RB, Keefe F. Race, Ethnicity and Pain. PAIN 2001;94(2):133–137.

[14] Fillingim RB. Individual differences in pain: understanding the mosaic that makes pain personal. Pain 2017;158 Suppl 1(Suppl 1):S11–S18.

[15] Franco M, Toomey T, DeBlaere C, Rice K. Identity incongruent discrimination, racial identity, and mental health for multiracial individuals. Counselling Psychology Quarterly 2021;34(1):87–108.

[16] Grol-Prokopczyk H. Sociodemographic disparities in chronic pain, based on 12-year longitudinal data. Pain 2017;158(2):313–322.

[17] Hamilton ER, Hale JM, Savinar R. Immigrant Legal Status and Health: Legal Status Disparities in Chronic Conditions and Musculoskeletal Pain Among Mexican-Born Farm Workers in the United States. Demography 2019;56(1):1–24.

[18] Hollingshead NA, Ashburn-Nardo L, Stewart JC, Hirsh AT. The Pain Experience of Hispanic Americans: A Critical Literature Review and Conceptual Model. The Journal of Pain 2016;17(5):513–528.

[19] Janevic MR, McLaughlin SJ, Heapy AA, Thacker C, Piette JD. Racial and Socioeconomic Disparities in Disabling Chronic Pain: Findings From the Health and Retirement Study. The Journal of Pain 2017;18(12):1459–1467.

[20] Jay MA, Bendayan R, Cooper R, Muthuri SG. Lifetime socioeconomic circumstances and chronic pain in later adulthood: findings from a British birth cohort study. BMJ Open 2019;9(3):1–10.

[21] Jimenez N, Garroutte E, Kundu A, Morales L, Buchwald D. A Review of the Experience, Epidemiology, and Management of Pain among American Indian, Alaska Native, and Aboriginal Canadian Peoples. The Journal of Pain 2011;12(5):511–522.

[22] Johannes CB, L. TK, Zhou X, Johnston JA, Dworkin RH. The Prevalence of Chronic Pain in United States Adults: Results of an Internet-Based Survey. The Journal of Pain 2010;11(11):1230–1239.

[23] Kim HJ, Yang GS, Greenspan JD, Downton KD, Griffith KA, Renn CL, Johantgen M, Dorsey SG. Racial and ethnic differences in experimental pain sensitivity: systematic review and meta-analysis. PAIN 2017;158(2):194–211.

[24] Kim HJ, Yang GS, Greenspan JD, Downton KD, Griffith KA, Renn CL, Johantgen M, Dorsey SG. Racial and ethnic differences in experimental pain sensitivity: systematic review and meta-analysis. PAIN 2017;158(2).

[25] Lavin R, Park J. A Characterization of Pain in Racially and Ethnically Diverse Older Adults: A Review of the Literature. Journal of Applied Gerontology 2012;33(3):258–290.

[26] Liu Y, Wheaton AG, Chapman DP, Cunningham TJ, Lu H, Croft JB. Prevalence of Healthy Sleep Duration among Adults — United States, 2014. MMWR 2016;65(6):137–141.

[27] Lokshin M. Semi-Parametric Difference-Based Estimation of Partial Linear Regression Models. Stata Journal 2006;6(3):377–383.

[28] McGuire TG, Miranda J. New Evidence Regarding Racial And Ethnic Disparities In Mental Health: Policy Implications. Health Affairs 2008;27(2):393–403.

[29] Meghani SH, Chittams J. Controlling for Socioeconomic Status in Pain Disparities Research: All-Else-Equal Analysis When “All Else” Is Not Equal. Pain Medicine 2015;16(12):2222–2225.

[30] Nahin RL. Estimates of Pain Prevalence and Severity in Adults: United States, 2012. The Journal of Pain 2015;16(8):769–780.

[31] Nahin RL. Pain Prevalence, Chronicity and Impact Within Subpopulations Based on Both Hispanic Ancestry and Race: United States, 2010-2017. The Journal of Pain 2021;Available online 23 February 2021.

[32] Ostrom C, Bair E, Maixner W, Dubner R, Fillingim RB, Ohrbach R, Slade GD, Greenspan JD. Demographic Predictors of Pain Sensitivity: Results From the OPPERA Study. The Journal of Pain 2017;18(3):295–307.

[33] Pitcher MH, Von Korff M, Bushnell MC, Porter L. Prevalence and Profile of High-Impact Chronic Pain in the United States. The Journal of Pain 2019;20(2):146–160.

[34] Portenoy RK, Ugarte C, Fuller I, Haas G. Population-based survey of pain in the united states: Differences among white, african american, and hispanic subjects. The journal of pain 2004;5(6):317–328.

[35] Quiton RL, Leibel DK, Boyd EL, Waldstein SR, Evans MK, Zonderman AB. Sociodemographic patterns of pain in an urban community sample: an examination of intersectional effects of sex, race, age, and poverty status. PAIN 2020;161(5).

[36] Rao JNK, Scott AJ. On Chi-Squared Tests for Multiway Contingency Tables with Cell Proportions Estimated from Survey Data. Annals of Statistics 1984;12(1):46–60.

[37] Raphael KG, Marbach JJ. When Did Your Pain Start?: Reliability of Self-Reported Age of Onset of Facial Pain. The Clinical Journal of Pain 1997;13(4):352–359.

[38] Reyes-Gibby CC, Aday LA, Todd KH, Cleeland CS, Anderson KO. Pain in Aging Community-Dwelling Adults in the United States: Non-Hispanic Whites, Non-Hispanic Blacks, and Hispanics. The Journal of Pain 2007;8(1):75–84.

[39] Rhudy JL, Huber F, Kuhn BL, Lannon EW, Palit S, Payne MF, Hellman N, Sturycz CA, Güereca YM, Toledo TA, Demuth MJ, Hahn BJ, Shadlow JO. Pain-related anxiety promotes pronociceptive processes in Native Americans: bootstrapped mediation analyses from the Oklahoma Study of Native American Pain Risk. Pain Rep 2020;5(1):e808–e808.

[40] Rhudy JL, Lannon EW, Kuhn BL, Palit S, Payne MF, Sturycz CA, Hellman N, Güereca YM, Toledo TA, Huber F, Demuth MJ, Hahn BJ, Chaney JM, Shadlow JO. Assessing peripheral fibers, pain sensitivity, central sensitization, and descending inhibition in Native Americans: main findings from the Oklahoma Study of Native American Pain Risk. PAIN 2020;161(2):388–404.

[41] Rice ASC, Smith BH, Blyth FM. Pain and the global burden of disease. Pain 2016;157(4):791–796.

[42] Riskowski JL. Associations of Socioeconomic Position and Pain Prevalence in the United States: Findings from the National Health and Nutrition Examination Survey. Pain Medicine 2014;15(9):1508–1521.

[43] Ross E, Huber F, Kuhn B, Lannon E, Sturycz C, Payne M, Hellman N, Toledo T, Güereca Y, Palit S, Demuth M, Shadlow J, Rhudy J. Assessing Chronic Pain Onset in Native Americans: Follow-Up Results from the Oklahoma Study of Native American Pain Risk (OK-SNAP). The Journal of Pain 2019;20(4, Supplement):S33.

[44] Ruggles S, Flood S, Foster S, Goeken R, Pacas J, Schouweiler M, Sobek M. IPUMS USA: Version 11.0 [dataset]. In: IPUMS editor, Vol. 2020. Minneapolis, MN: IPUMS. org, 2021.

[45] Steingrímsdóttir óA, Landmark T, Macfarlane GJ, Nielsen CS. Defining chronic pain in epidemiological studies: a systematic review and meta-analysis. PAIN 2017;158(11):2092–2107.

[46] Turner TM, Luea H. Homeownership, wealth accumulation and income status. Journal of Housing Economics 2009;18(2):104–114.

[47] U.S. Census Bureau. Projected Race and Hispanic Origin: Main Projections Series for the United States, 2017-2060. Washington DC: U.S. Census Bureau, Population Division, 2018.

[48] U.S. Census Bureau. Annual Estimates of the Resident Population by Sex, Age, Race Alone of in Combination, and Hispanic Origin for the United States: April 1, 2010 to July 1, 2019 (NC-EST2019-ASR5H). Washington DC: US Census Bureau, Population Division, 2020.

[49] US Census Bureau. Selected Social Characteristics in the United States, Table DP02, 2021.

[50] Vaughn IA, Terry EL, Bartley EJ, Schaefer N, Fillingim RB. Racial-Ethnic Differences in Osteoarthritis Pain and Disability: A Meta-Analysis. The Journal of Pain 2019;20(6):629–644.

[51] Veenstra G. Mismatched racial identities, colourism, and health in Toronto and Vancouver. Social science & medicine (1982) 2011;73(8):1152–1162.

[52] Williams DR. Miles to Go before We Sleep: Racial Inequities in Health. Journal of Health and Social Behavior 2012;53(3):279–295.

[53] Yong RJ, Mullins PM, Bhattacharyya N. The prevalence of chronic pain among adults in the United States. PAIN 2021;Articles in Press.

[54] Zajacova A. Health in Working-Age Americans: Adults with a High School Equivalency (GED) Diploma Are Similar to Dropouts, Not High School Graduates. American Journal of Public Health 2012;102(S2):S284–S290.

[55] Zajacova A, Grol-Prokopczyk H, Zimmer Z. Pain Trends Among American Adults, 2002–2018: Patterns, Disparities, and Correlates Demography 2021;58(2):711–738.

[56] Zajacova A, Grol-Prokopczyk H, Zimmer Z. Sociology of Chronic Pain. Journal of Health and Social Behavior 2021;Forthcoming.

[57] Zajacova A, Rogers RG, Grodsky E, Grol-Prokopczyk H. The Relationship between Education and Pain among Adults Aged 30-49 in the United States. Journal of Pain 2020;21(11-12):1270–1280.

[58] Zelaya CE, Dahlhamer JM, Lucas JW, Connor EM. Chronic Pain and High-impact Chronic Pain Among U.S. Adults, 2019. NCHS Data Brief, Vol. 390. Hyattsville, MD: National Center for Health Statistics, 2020.

